# Deep Longitudinal Clusters of Type 2 Diabetes Pathophysiology and their Risk of Cardiovascular Disease Events and All-Cause Mortality

**DOI:** 10.64898/2026.06.01.26354645

**Authors:** Jithin Sam Varghese, Jiali Guo, Daniel Hua, Theodore Hung, Zhongyu Li, Shengpu Tang, Shivani A. Patel, Joyce C. Ho

**Author notes:** Corresponding Author:* Jithin Sam Varghese, Hubert Department of Global Health, Rollins School of Public Health, Emory University, Atlanta, GA, 30322, USA.

## Abstract

**Objective:** Despite the complex and non-linear progression of diabetes, its shared pathways with atherosclerotic cardiovascular disease (ASCVD) are conventionally described using models based on single time points. We identified longitudinal diabetes clusters before diagnosis using deep learning and studied their association with ASCVD events and mortality.

**Methods:** We analyzed 157,670 visits from 15,871 adults (25-65 years) without diabetes from four pooled U.S. cohorts (median follow-up: 22 years [IQR: 9-30]). A gated recurrent unit model with decay (GRU-D) was used to predict 1-year risk of diabetes or censoring within 10 years, by learning longitudinal embeddings across 25 clinical characteristics and biomarkers. Parallel Factor Analysis-2 (PARAFAC-2) and Gaussian mixture models (GMM) were used to group longitudinal participant representations as clusters. Landmark time Cox proportional hazards regressions, relative to last observation in the training window, were used to study covariate-adjusted associations of clusters with ASCVD and mortality. Prognostic utility of clusters beyond the PREVENT risk score was assessed using Harrell’s C-index. Findings were replicated in a fifth cohort.

**Results:** The analytic sample was aged 49 years [SD: 11], 58% female, and 68% white; 1,202 (8%) developed diabetes within the first 10 years. We identified five clusters (Cluster A to E) that differed in their clinical characteristics over time. Cluster E (46%) had the highest cumulative incidence of diabetes in the study period, followed by Cluster C (40%) and Cluster A (38%). Cluster C, which was defined by older age, high blood pressure, and suboptimal renal function at the first visit, had higher rates of ASCVD (HR: 1.09, 95%CI: 0.98-1.21) and mortality (HR: 1.08, 95%CI: 1.00-1.16), relative to Cluster A despite being similar in age and BMI at the first visit. Relative to Cluster A, all other clusters had similar or lower rates of ASCVD and mortality. We observed substantial cluster effects for three clusters (Clusters C to E), which were based on only two cohorts. The two clusters (Clusters A and B) that included participants from all four cohorts were reproduced in the fifth cohort and showed similar rates of outcomes. Clusters did not improve ASCVD prognosis, relative to a model that included only the PREVENT risk score.

**Conclusions:** Longitudinal clusters reveal substantial heterogeneity in the period before diabetes diagnosis, and their risk for ASCVD and mortality. However, clusters discovered may, in part, be explained by cohort effects from variations in recruitment and visit patterns after recruitment.

**RESEARCH IN CONTEXT:** *Why did we undertake this study?:* Pathophysiological heterogeneity before diabetes diagnosis is known to be dynamic over time. Previous studies that captured pre-diagnosis heterogeneity were limited to single time-point biomarkers.

*What is the specific question we wanted to answer?:* Can we identify longitudinal clusters of the period before diabetes diagnosis in a pooled cohort of U.S. adults (25-65 years) without diabetes using a deep learning-enabled workflow and does cluster membership predict risk of future ASCVD events and mortality?

*What did we find?:* Individuals without diabetes belonged to five longitudinal clusters with heterogeneous biomarker trajectories. The clusters showed clinically meaningful differences in relative incidence of ASCVD events and mortality.

*What are the implications of our findings?:* Risk stratification based on longitudinal trajectories of biomarkers can inform precision prevention of diabetes, ASCVD events, and premature mortality.

## INTRODUCTION

Type 2 diabetes is a heterogeneous disease in its clinical presentation and response to treatment (1,2). Precision medicine aims to classify individuals with type 2 diabetes into subgroups based on etiological and pathophysiological risk factors, variations in treatment response, and risk for diabetes complications (3–5). Incorporating diabetes heterogeneity in clinical care can move us away from a one-size-fits-all paradigm for disease treatment and towards tailored management strategies (6,7).

Data-driven clustering is a useful approach to understand disease heterogeneity and a prognostic tool for evaluating the risk of diabetes and its complications (3). Yet, current approaches to characterize heterogeneity in the period before diabetes diagnosis have several shortfalls. First, most studies have relied on biomarkers measured at a single time point (4,8). Single measures are inadequate to capture trajectories in the period preceding diabetes diagnosis, which have been shown to be non-linear (9,10). Second, traditional clustering methods are largely applied to complete cases and thus omit individuals with missing data on one or more biomarkers. Such omission unintentionally induces biases in the resulting clusters (8,11–13) and does not reflect real-world practice where not all individuals may have all biomarkers available. Third, cluster discovery and risk modeling for diabetes are separate steps and therefore may not capture the true underlying pathophysiology of diabetes heterogeneity (8).

Deep learning can overcome these limitations of traditional clustering approaches (14,15). One approach that combines representation of longitudinal patient data with varying follow-up, missing data imputation, and risk prediction is the Gated Recurrent Unit with Decay (GRU-D) (16). By training a GRU-D on longitudinal data of individuals without diabetes from diverse population-based cohorts, our primary objective is to learn longitudinal representations of clinical variables and biomarkers as neural network embeddings. We then clustered the embeddings using tensor decomposition and Gaussian Mixture Models to study their time to atherosclerotic cardiovascular disease (ASCVD) events and death. Findings from this study can inform the development of novel risk stratification tools for diabetes and its complications that incorporate irregular, real-world longitudinal data.

## METHODS

### Study Population

We pooled data from four prospective community-based studies in the United States: the Atherosclerosis Risk in Communities (ARIC) Study, Coronary Artery Risk Development in Young Adults (CARDIA) Study, Diabetes Prevention Program Trial and Outcomes Study (DPP/DPPOS), and Jackson Heart Study (JHS). ARIC is a cohort of adults from four communities to understand risk factors for cardiovascular and cognitive health. CARDIA enrolled young adults from four cities to study the lifestyle, socioeconomic and psychosocial risk factors for ASCVD. DPP was a randomized trial of individuals at high risk of type 2 diabetes. Individuals were assigned to one of three intervention arms (metformin, lifestyle, troglitazone [later discontinued]) or a control arm. DPPOS was the follow-up of DPP to evaluate the sustained benefits of diabetes prevention; we restricted our analysis of DPPOS to individuals from the metformin, lifestyle, and control arms. JHS is a prospective study of Black adults from one city to identify genetic and environmental risk factors of ASCVD. We excluded JHS participants who were in ARIC. The Multi-Ethnic Study of Atherosclerosis (MESA) cohort was used to validate the findings and consists of individuals from six cities who were prospectively followed to understand the role of subclinical disease and other risk factors of ASCVD.

We restricted our analysis to those without diabetes aged 25 to 65 years at the start of follow-up, with at least two visits before diabetes diagnosis or censoring in the first ten years, and at least one subsequent visit where age of diabetes diagnosis could be reliably ascertained (e.g., visit 4 of ARIC). Recruitment strategy and visit information for each cohort are presented in *Supplementary Table 1 and Supplementary Table 2*. The analysis framework and the participant flowchart are presented in *Supplementary Figure 1* and *Supplementary Figure 2*.

### Data collection and variable specification

We extracted and harmonized sociodemographic and clinical data across studies. Sociodemographic data included age, sex (male, female), and race and ethnicity. We harmonized cohort-specific race classifications (without differentiated ethnicity) as four categories: Hispanic, White, Black, and Other. Similarly, we harmonized clinical history and assessments to derive common measures for smoking (never, former, current), alcohol use (never, former, current), medications (high blood pressure, lipid lowering medication), anthropometry (height, BMI, systolic and diastolic blood pressure, waist circumference) and routinely collected laboratory markers for monitoring dysglycemia (HbA1c, fasting glucose, fasting insulin), lipids (total cholesterol, LDL cholesterol, HDL cholesterol, triglycerides), blood-based renal function (serum creatinine). Homeostatic indices for beta-cell function and insulin resistance were calculated using the Oxford HOMA2 calculator. Although risk enhancers (Apo[a], Apo[b]), urinary biomarkers, and liver function tests were available for some cohorts, these were largely missing across most study visits and hence not included. A summary of measurement protocols for different variables are presented in *Supplementary Table 3*.

Diabetes was determined based on self-reported diagnosis, a diagnosis confirmed by a physician or healthcare provider, or reported use of diabetes medications within the preceding year, or elevated laboratory biomarkers at the study exam (HbA1c ≥6.5% or fasting plasma glucose (FPG) ≥126 mg/dL, or a 2-hour glucose ≥200 mg/dL following an oral glucose tolerance test). ASCVD was defined as one of coronary heart disease [CHD], myocardial infarction, or stroke (17). All-cause mortality was adjudicated using study-specific protocols. Time to all events were determined based on cohort-specific definitions (*Supplementary Table 3*).

### Statistical Analysis

A detailed description of the analytical workflow is provided in the *Supplementary Methods Note*.

#### Longitudinal Representations using Deep Learning

The longitudinal pooled cohort data included data from all available visits, from the first study visit without diabetes up to ten years of follow-up or onset of diabetes, whichever came first. We chose a ten year period for accumulating cases of diabetes and non-linear changes in biomarkers. We included a maximum of one visit (observation) per participant per year. We used the median value of biomarkers for participants from DPP/OS with more than one visit in a year.

We used a GRU-D, a type of recurrent neural network to study the risk of diabetes in the subsequent year for each of the ten years included (16). The GRU-D additionally takes in two inputs to form a fixed-size tensor: the masking vector and the time interval vector for combinations of each individual, time point, and variable. The masking vector is a sequence of binary (1/0) variables based on variable availability (1 = available, 0 = missing data). The time interval vector is calculated relative to the last available observation of the variable. To handle missing data, GRU-D applies a decay rate based on the time interval vector separately for each variable and the hidden nodes. The embeddings in the hidden layer of the GRU-D are updated at each iteration based on a combination of observed data, masking vector, and time decay to minimize the difference between the actual status and predicted probability for 1-year risk of diabetes. This modeling framework is analogous to a pooled logistic regression for survival analysis that can incorporate time varying covariates but without pre-specifying non-linear transformations of variables and their statistical interactions.

Categorical variables were transformed into dummy variables (one-hot encoding) and continuous variables were standardized based on the pooled mean and standard deviation. The model was fit on 80% of the analytic sample (n = 12,696) using 30 input nodes and a single layer of 24 hidden nodes with standard GRU gates for reset and update with 20% dropout, learning rate of 10^-3^ and weight decay of 10^-5^ (16). The parameters were chosen based on simulation studies for datasets of similar dimensions (16). Given the imbalanced nature of the outcome, model performance was evaluated by average area under receiver operating characteristic curve (AUROC), area under precision-recall curve (AUPRC) and Brier scores for the 1-year risk of diabetes for each year using a held-out sample (n = 3,175 or 20%) (18). The final three-dimensional tensor of GRU-D embeddings (individuals, years, hidden nodes) were extracted for clustering.

#### Tensor Decomposition

We clustered the GRU-D embeddings by fitting a parallel factor analysis model-2 (PARAFAC-2) using the alternating least squares algorithm (19). PARAFAC-2 is an unsupervised algorithm that decomposes tensors into three matrices of individuals, time, and variables across latent components, analogous to a principal components analysis for matrices. We identified the optimal number of components (range: 2 to 8) as six based on the scree plot of the proportion of variance in the original tensor of GRU-D embeddings explained by the reconstruction. An individual’s loadings on each component were extracted from the model.

#### Defining Longitudinal Clusters

Individual loadings were clustered using Gaussian Mixture Models (20). The optimal number of clusters (range: 2 to 10) was identified as five based on the largest value of the median adjusted Rand Index (ARI) > 0.8 over 200 bootstrap samples. Individuals were assigned to their most probable cluster for studying their associations with outcomes.

#### Survival Analysis

To study the association of clusters with ASCVD and mortality, we conducted a landmark survival analysis after diabetes incidence or censoring at the end of the training window (i.e., 10 years) using Cox proportional hazards regression. Participants, without an ASCVD event before or during the training window, were followed till the outcome of interest (median; ASCVD: 22 years; mortality: 28 years) or censoring (last follow-up or death). We adjusted for age at landmark time, sex, and race & ethnicity to study the associations of cluster membership (hazard ratios and 95% robust confidence intervals) with ASCVD and mortality separately (21). Next, we compared the clusters to the Predicting Risk of Cardiovascular Disease EVENTs (PREVENT-base) 10-year risk scores (17), calculated at the end of the training window using Harrell’s c-index.

To inform our understanding of disease progression, univariate biomarker trajectories of the clusters, without covariate adjustment, were generated from generalized linear mixed effects models with natural cubic splines of time relative to the first study visit (22). We visualized trajectories recognizing that the true latent representations are more complex and non-linear with interactions between different biomarkers.

To evaluate the replicability of these clusters, we implemented the GRU-D and clustering workflow in the held-out validation cohort. Analyses were carried out using Python 3.11.4 with tensorflow and R 4.5.0 with multiway, mclust, survival and lme4 packages.

## RESULTS

The pooled analytic cohort consisted of 15,871 individuals without physician-diagnosed diabetes at the first visit. Participants were aged 48.7 years (SD: 10.9), 57.7% female, 68.5% NH White, 27.8% NH Black, 2.8% Hispanic, and 0.9% from other racial groups. Majority of the participants belonged to ARIC (63%). Most participants across cohorts had 2 visits [IQR: 2-4] in the first ten years of follow-up, with participants in the DPP/OS study having more frequent visits than other cohorts (*Supplementary Figure 3, Supplementary Figure 4*). Rates of missingness across variables over the ten-year period are presented in *Supplementary Figure 5*.

During the ten-year training window with 50,359 observations, 1,202 individuals developed diabetes in the subsequent year. The GRU-D model showed an average AUROC of 0.90, AUPRC of 0.28, and Brier scores of 0.01 in the held-out sample over the training window for 1-year risk of diabetes (*Supplementary Figure 6*). Longitudinal embeddings were then used to identify five clusters using tensor decomposition and Gaussian Mixture Models for clustering (*Supplementary Figure 7*).

Clusters differed in their socio-demographic and clinical characteristics at the first visit, and over time. Cluster C (21%) participants were aged 54.6 years (SD: 5.8), most likely to be male (51%), had higher SBP (143.5 mmHg), and were more likely to use antihypertensives (35%) and lipid lowering medications (32.9%) at the first visit (*Table 1*). Cluster B (19%) participants were younger (33.9 years [SD: 10.4]) and more likely to be Black (54%) relative to the other clusters. Cluster A (25%) participants were aged 49.8 years (SD: 9.7), high BMI (30.1 kg/m^2^) and SBP (133.6 mmHg). Cluster D (27%) participants were majority White (90%) and had the lowest BMI (24.9 kg/m^2^) relative to other clusters. Despite the lower BMI relative to Cluster B (26 kg/m^2^), Cluster D participants had a higher waist circumference (89.6 cm vs 83.2 cm for Cluster B). Cluster E (8%) participants were aged 48.9 years (SD: 8.3), most likely to be female (69%) and Hispanic (17%). Relative to other clusters, Cluster E participants had the highest BMI (34.2 kg/m^2^), waist circumference (102.5 cm), and triglycerides (159.4 mg/dL) and lowest HDL (45.7 mg/dL). Clusters also showed varied cohort composition, with Cluster C and Cluster D enriched for ARIC, and Cluster E consisting mostly of DPP/OS.

**Table 1.**
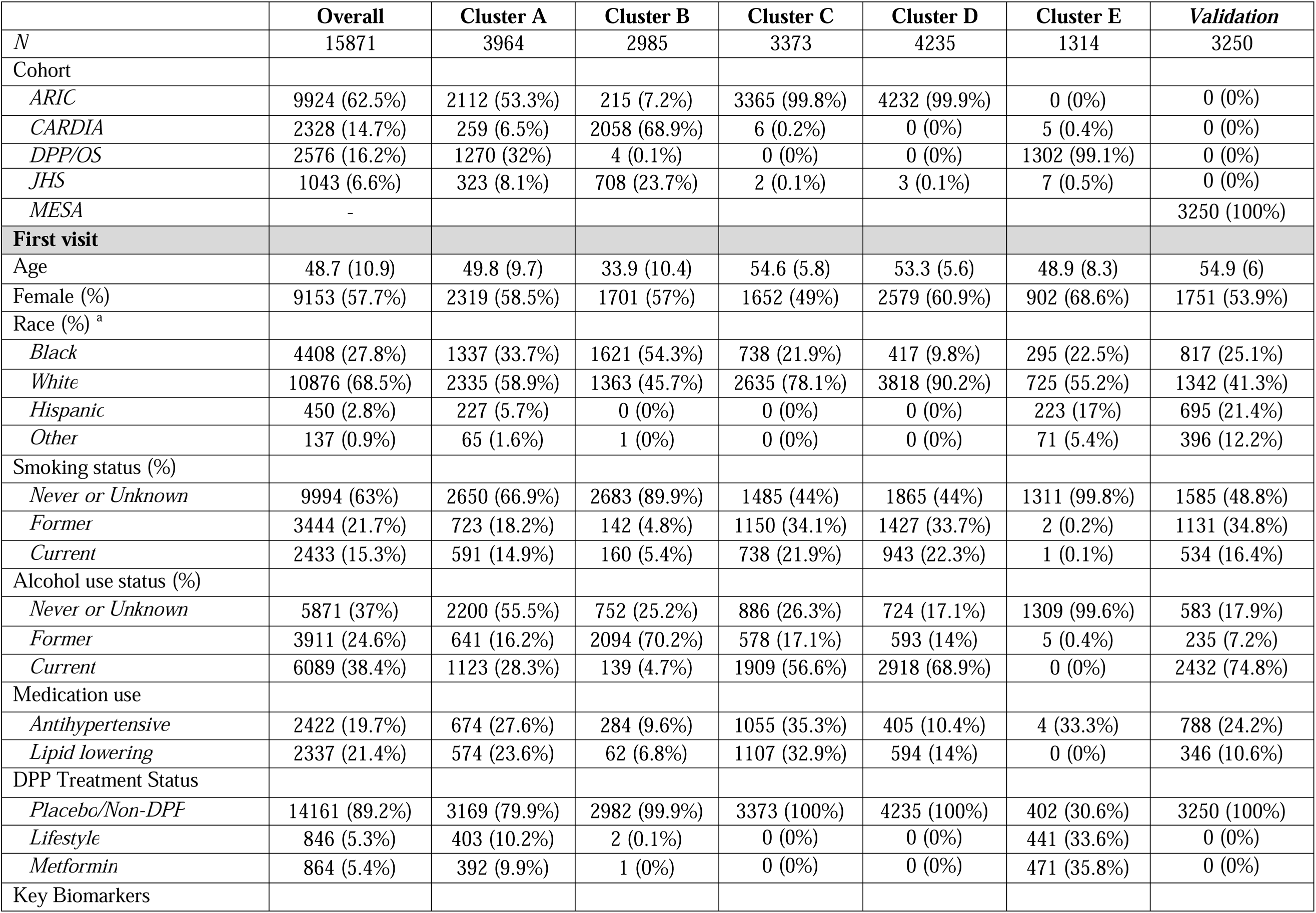
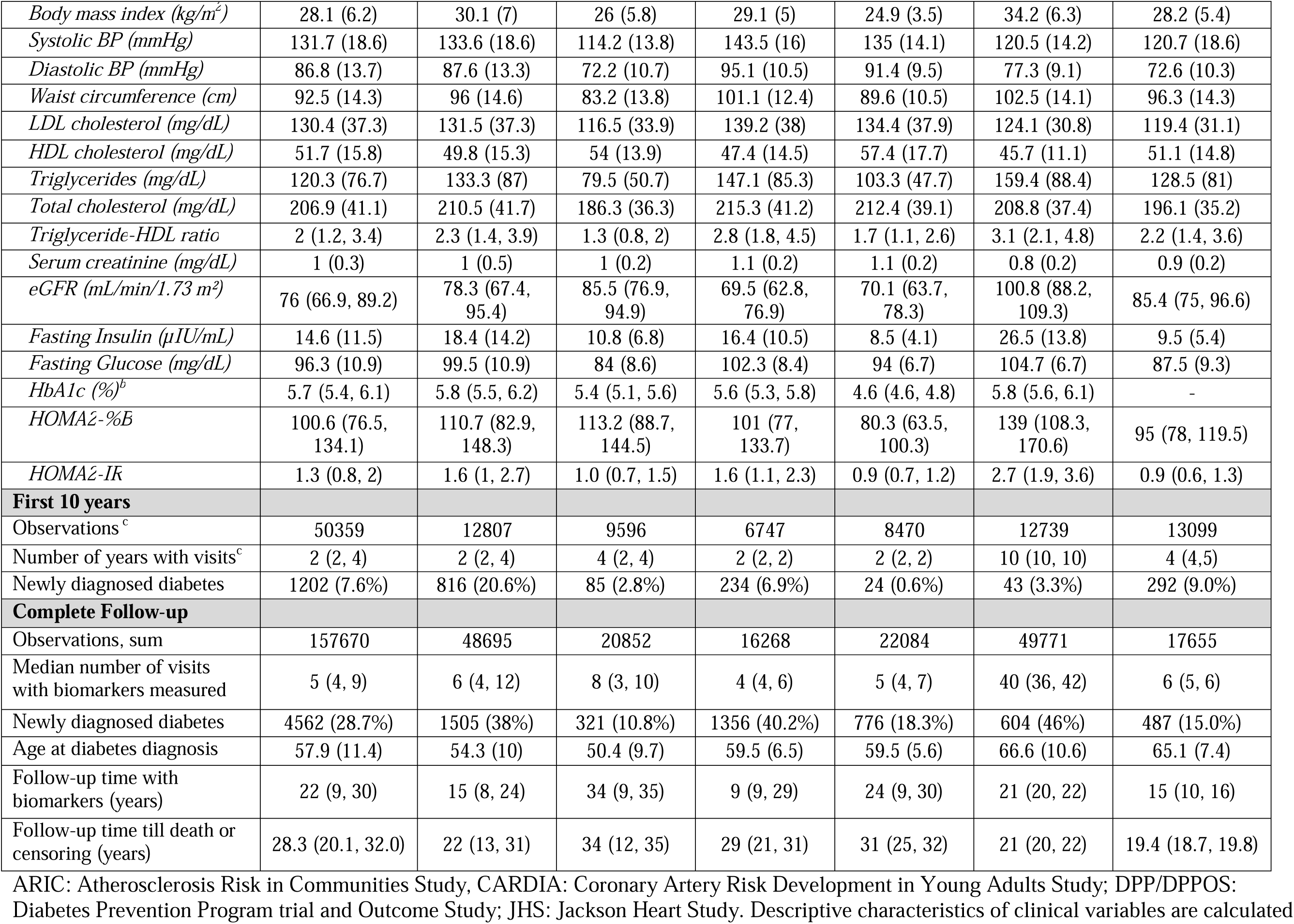
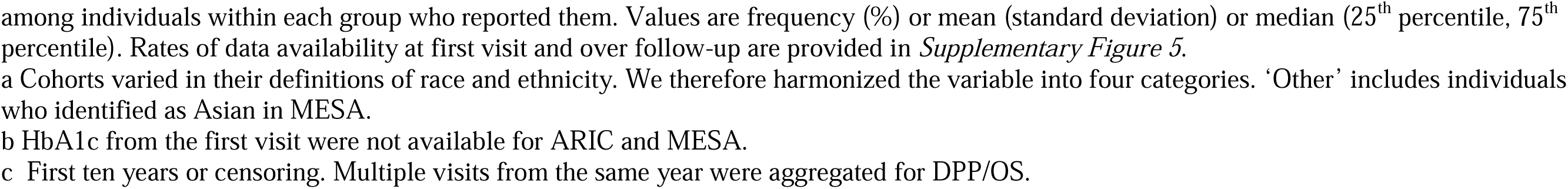
Descriptive characteristics at first visit by longitudinal clusters of individuals without diabetes in pooled cohorts.

Cumulative incidence of ASCVD varied between clusters (*Figure 1*). After the landmark time, relative to Cluster A, Cluster C (HR: 1.29 [95%CI: 1.17-1.43]) had higher rates, while Cluster D (HR: 0.91 [95%CI: 0.82-1.00]), Cluster E (0.64 [95%CI: 0.51-0.80]) and Cluster B (0.50 [95%CI: 0.42-0.59]) had lower rates. After adjusting for age, sex, and race, Cluster C continued to have higher rates (HR: 1.09 [95%CI: 0.98-1.21]) and other clusters had lower rates (*Table 2*).

**Figure 1.**
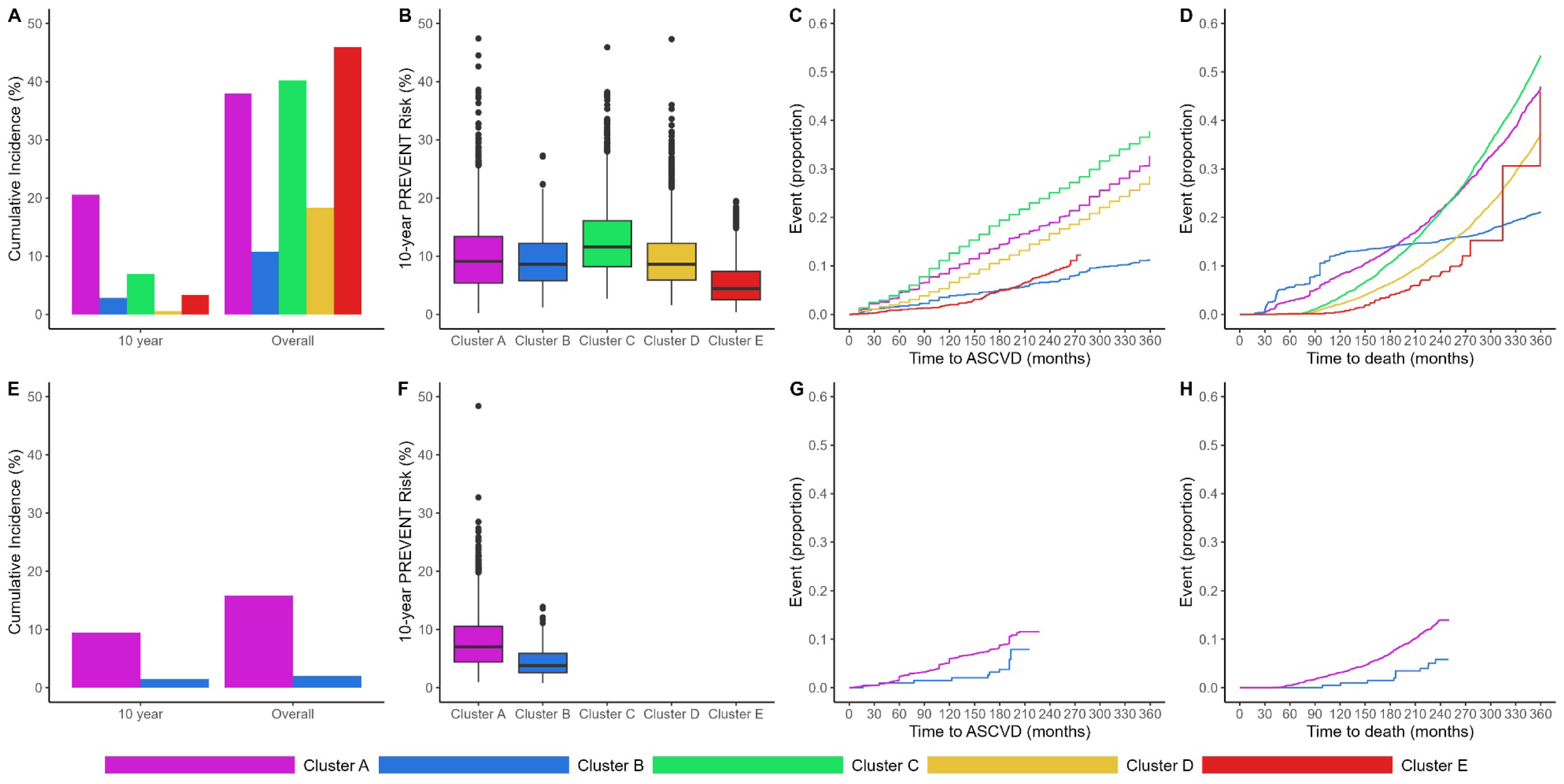
Time to cardiometabolic events and mortality relative to first visit. Crude cumulative incidence curves (and 95% confidence intervals) for time to event. Top row is the cumulative incidence of diabetes (Panel A), distribution of the PREVENT scores at the last visit in the 10-year training window (Panel B), time to ASCVD (Panel C; myocardial infarction, coronary heart disease, or stroke), and time to death (Panel D) for the pooled cohort sample. Bottom row is the same set of figures (Panels E-H) for the validation cohort.

**Table 2.** Relative hazards of longitudinal clusters with atherosclerotic cardiovascular events and mortality in pooled cohorts.

|  |  | *Atherosclerotic cardiovascular disease events* |  |  |  | *All-cause mortality* |  |  |
| --- | --- | --- | --- | --- | --- | --- | --- | --- |
|  | Follow-up (months) | Events/N | Unadjusted HR | Adjusted HR | Follow-up (months) | Events/N | Unadjusted HR | Adjusted HR |
| ***Pooled*** |  |  |  |  |  |  |  |  |
| Overall | 228 (0-369) | 2722/12625 | - | - | 283 (0-383) | 5699/14454 | - | - |
| Cluster A | 203 (0-369) | 610/2971 | Ref (1.00) | Ref (1.00) | 236 (0-383) | 1324/3285 | Ref (1.00) | Ref (1.00) |
| Cluster B | 285 (3-359) | 164/1151 | 0.50 (0.42, 0.59) | 0.80 (0.62, 1.04) | 299 (0-383) | 319/2271 | 0.24 (0.21, 0.27) | 0.88 (0.73, 1.05) |
| Cluster C | 228 (12-368) | 904/3146 | 1.29 (1.17, 1.43) | 1.09 (0.98, 1.21) | 271 (0-371) | 2048/3369 | 1.33 (1.24, 1.43) | 1.08 (1.00, 1.16) |
| Cluster D | 275 (12-311) | 955/4084 | 0.91 (0.82, 1.00) | 0.85 (0.76, 0.94) | 299 (2-335) | 1893/4230 | 0.83 (0.77, 0.89) | 0.81 (0.75, 0.87) |
| Cluster E | 132 (1-263) | 89/1273 | 0.64 (0.51, 0.8) | 0.61 (0.47, 0.78) | 132 (1-311) | 115/1299 | 0.80 (0.66, 0.97) | 0.84 (0.67, 1.05) |
| ***Validation*** |  |  |  |  |  |  |  |  |
| Overall | 72 (1-168) | 105/2238 | - | - | 115 (0-204) | 390/3240 | - | - |
| Cluster A | 72 (1-168) | 97/2060 | Ref (1.00) | Ref (1.00) | 115 (0-204) | 379/3037 | Ref (1.00) | Ref (1.00) |
| Cluster B | 72 (3-144) | 8/178 | 0.92 (0.45, 1.88) | 1.07 (0.51, 2.26) | 116 (27-193) | 11/203 | 0.44 (0.24, 0.81) | 0.57 (0.31, 1.05) |

Hazard ratios from Cox proportional hazards regression from diabetes incidence (landmark time) or censoring in the 10-year training window till end of follow-up in months (min-max), adjusted for sex and race. We stratified by age at landmark time. We excluded individuals who experienced any ASCVD prior to landmark time or with missing data on the outcome. We did not adjust for study fixed effects since three clusters included mostly a single cohort. Clusters 1, 3, and 5 were not identified in the validation sample.

Differences between clusters in cumulative incidence of mortality were less clear (*Figure 1*). After landmark time, and adjusting for covariates, relative to Cluster A, Cluster C (HR: 1.08 [95%CI: 1.00-1.16]) had higher rates of mortality, while Cluster D (HR: 0.81 [95%CI: 0.75-0.87]), Cluster B (HR: 0.88 [95% CI: 0.73-1.05]), and Cluster E (HR: 0.84 [95%CI: 0.67-1.05]) had lower rates.

In the subsample for whom all variables were available to calculate PREVENT 10-year risk, a model with clusters demonstrated no incremental value for predicting ASCVD (n = 10,109; C-index: 0.63 [95%CI: 0.62-0.64]) and mortality (n = 10,609; C-index: 0.69 [95%CI: 0.68-0.70]) relative to a model without clusters (ASCVD: 0.64, mortality: 0. 69).

Trajectories of biomarkers also varied in their shape (baseline, trend) between clusters (*Figure 2*). Cluster A displayed increasing waist circumference, and declining eGFR and HOMA2-%B. Cluster B displayed trajectories of increasing BMI and fasting glucose, with normal SBP and eGFR. Cluster C displayed stable BMI, persistently high SBP, and low eGFR relative to other clusters. Cluster D displayed metabolically healthy trajectories across all biomarkers, except for the declining HDL at later stages. Cluster E displayed a trajectory of decreasing BMI from very high baseline, HOMA2-%B and eGFR, and increasing SBP.

**Figure 2.**
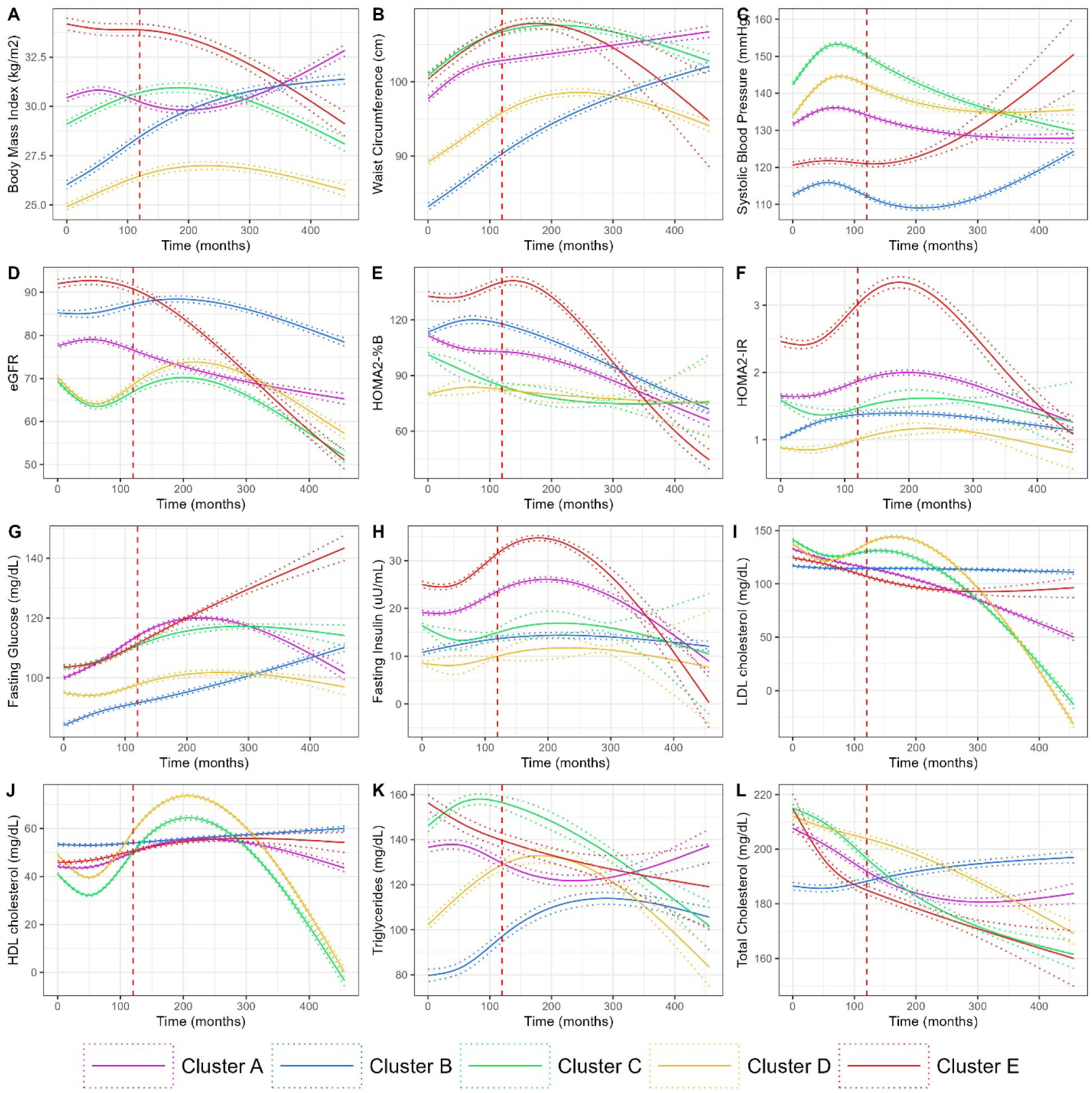
Modeled trajectories of clinical characteristics among clusters in pooled cohorts. Unadjusted univariate trajectories (and 95% confidence intervals) from linear mixed effects models (n = 15,871 individuals) for routine vitals and labs. Person level trajectories are provided in *Supplementary File 2*. Red vertical dashed line indicates the maximum training window (10 years).

The validation cohort (MESA; n = 3250) was older (54.9 years [SD: 6]) and ethnically diverse, with similar BMI but lower SBP and LDL relative to the training sample (*Table 1*). Average follow-up was shorter in the validation sample (19.4 years), compared to the training sample (28.3 years). During the 10-year training window, 292 individuals developed diabetes. The GRU-D model had an average AUROC of 0.83, AUPRC of 0.12, and Brier score of 0.01 over the training window for the validation sample (*Supplementary Figure 6*). Majority of the sample was classified as Cluster A (94%), followed by Cluster B (6%) (*Supplementary Table 4*). Crude rates of ASCVD and mortality mirrored those of the pooled training sample (*Figure 1*). After the landmark time, and adjusting for covariates, relative to Cluster A, Cluster B was not associated with ASCVD events (HR: 1.11 [95%CI: 0.53-2.32]) but may have lower rates of mortality (HR: 0.57 [95%CI: 0.31-1.05]).

## DISCUSSION

Using deep learning to characterize the time-varying risk of diabetes over a ten-year window, we identified heterogeneous longitudinal clusters using 25 clinical variables in the period before diabetes diagnosis. The model showed good discriminative performance and accuracy over time. A high-risk cluster (Cluster C) defined by older age, high blood pressure, and suboptimal renal function had higher rates of ASCVD and mortality, relative to other clusters, including those of similar age and higher BMI at baseline. Two clusters (Cluster A and Cluster B) were identified in an independent validation cohort and demonstrated similar crude rates of outcomes. However, the clusters did not demonstrate additional predictive utility for ASCVD events and mortality compared to the PREVENT 10-year risk at the last available visit. Nevertheless, we observed substantial cohort effects for Clusters C, D, and E that may have been influenced by the visit patterns and consequently, missing data associated with different cohorts.

The period before diabetes diagnosis is known to be a complex longitudinal process marked by heterogeneity in β-cell function, insulin resistance, and other pathophysiological processes (23). For instance, participants in the Whitehall II study in the UK and pooled cohorts from the USA displayed non-linear trajectories in fasting glucose and insulin, especially in the five years before diabetes onset (9,10). However, most data-driven subtyping studies of the period before diabetes were limited to a single time point, and clustered biomarkers from individuals without diabetes into discrete subtypes (24,25). Other studies leveraged polygenic risk scores to characterize genetic and phenotypic heterogeneity for risk stratification (4,26). For instance, six heterogeneous subtypes were identified through data-driven subtyping of 899 participants of the TUEF/TULIP study and replicated among 6,810 participants of the Whitehall II study (8). Three of these subtypes were at elevated risk of progression to diabetes, displayed differences in fat accumulation and β-cell function, and were at elevated risk of diabetes complications. A recent study characterized longitudinal metabolic trajectories in the DPP/OS using tensor decomposition and identified distinct subgroups of prediabetes, with varying susceptibility for microvascular and macrovascular complications (27). Our findings add to this growing literature, by leveraging longitudinal data for risk stratification for diabetes and its complications.

We observed substantial cohort effects in clustering, with three clusters consisting of ARIC (Cluster C, Cluster D) and DPP/OS (Cluster E). ARIC was the largest community cohort included, and participants substantially varied in their rates of diabetes and ASCVD. DPP/OS was a trial of participants at high risk of diabetes with strict eligibility criteria (overweight with both impaired fasting glucose and impaired glucose tolerance) and frequent follow-ups (28). The two clusters that were reproduced in the validation sample included participants from all cohorts, suggesting that the workflow can be generalized to capture longitudinal trajectories among participants with varying follow-up intervals and measurements.

The GRU-D model enabled accurate assessment of 1-year risk of diabetes in the training window and identified clusters prognostic of ASCVD events and all-cause mortality. By identifying clusters representing the time-varying risk of diabetes and evaluating their association with complications, we can derive novel insights into shared pathophysiology of these conditions. Cluster C was identified as a high risk cluster for ASCVD and mortality, despite lower rates of diabetes, relative to Cluster A. Furthermore, Cluster E was of similar age as Cluster C but had higher BMI, fasting glucose, and HbA1c at baseline. The elevated rates of ASCVD among Cluster C may be explained by the persistently higher systolic BP and LDL cholesterol over time. Nevertheless, the suboptimal performance of clusters for ASCVD risk stratification, relative to PREVENT, was not surprising and may be explained by differences in training objectives. The clusters were based on GRU-D embeddings trained to predict the diagnosis of diabetes in a 10-year training window, while PREVENT risk scores calculate the subsequent 10-year risk of ASCVD based on the last time point in that 10-year window. A study using the ADOPT and RECORD trials also showed that risk scores based on continuous clinical features were more predictive of glycemic response, compared to clusters of newly diagnosed diabetes (29).

This study has several strengths such as the large and diverse sample, using supervised learning to learn longitudinal representations that are associated with onset of diabetes, and long duration of follow-up for ASCVD and mortality. Nevertheless, there are some weaknesses. First, some studies were initiated at a time when HbA1c was not available (e.g., in ARIC) or used as a diagnostic tool (e.g., DPP/OS). This may have resulted in individuals who may have otherwise been diagnosed with diabetes based on current practice being included in the analytic sample. Furthermore, we were unable to distinguish between type 2 diabetes and other less common forms of adult-onset diabetes (e.g., Latent Autoimmune Diabetes in Adults). Second, although GRU-D and other deep representation learning approaches simultaneously learn missingness patterns and predict time to event, they operate under a missing at random assumption, such that available data is informative of missing values of different biomarkers (14,16,30). Furthermore, by limiting our sample to those with at least two visits, we may be restricting to engaged participants of different studies. Third, deep learning algorithms, by design, are inherently uninterpretable and cluster labels are based on post-hoc trajectories from statistical models (31,32). For instance, we used natural cubic splines to study differences between clusters. However, the true latent representations may be more complex, non-linear trajectories that incorporate interactions between different biomarkers. Nevertheless, approximations such as ours are informative for explaining the underlying disease progression and interpreting clusters.

In summary, longitudinal clusters of clinical variables that incorporate irregular, real-world data can be prognostic of diabetes and its complications. However, these clusters did not improve risk stratification for ASCVD and mortality beyond an established risk score. Future studies should evaluate if supervised approaches that can simultaneously cluster and predict risk of outcomes, and incorporating additional modalities such as continuous glucose monitoring, genomics, and data from electronic health records can improve risk stratification among individuals without diabetes.

## Abbreviations

ARIC: Atherosclerosis Risk in Communities
CARDIA: Cardiovascular Risk in Young Adults
GRU-D: Gated Recurrent Unit-Decay
JHS: Jackson Heart Study
RNN: Recurrent Neural Networks

## ACKNOWLEDGEMENTS

Ethics approval and consent to participate

We were exempt from ethical approval for the analysis of secondary datasets. All participants of cohort studies and trials gave written informed consent before participation.

## Funding and Assistance

None

## Author contributions

JSV conceptualized the study with inputs from JCH. JG, DH, & TH led the data extraction. JSV led the analysis and wrote the first draft. JSV and JG are the guarantors of this work, and as such, had full access to all the data in the study and take responsibility for the integrity of the data and the accuracy of the data analysis. JG, DH, TH, ZL, ST, SAP, and JCH reviewed and edited the manuscript. All authors approved the final version of the manuscript.

## Personal Thanks

We thank the participants and study investigators of ARIC, CARDIA, DPP and Outcomes Study, JHS, and MESA for facilitating this study.

## Data availability

The code for the analysis will be made available upon publication at https://github.com/chroniq-lab/cohorts-gru-d. Data for the National Institutes of Health funded cohort studies are available for request from the NIDDK Central Repository (NIDDK-CR; https://repository.niddk.nih.gov) and NHLBI Biolincc (https://biolincc.nhlbi.nih.gov/home/) to registered users.

