## Supplementary Material for "Deep Longitudinal Clusters of Type 2 Diabetes Pathophysiology and their Risk of Cardiovascular Disease Events and All-Cause Mortality"

### Supplementary Table 1. Sources and procedures for measurement of key variables

| **Cohort** | **Active Years** | **Study Type** | **Description** | **Retention** |
| --- | --- | --- | --- | --- |
| Atherosclerosis Risk in Communities (ARIC)  https://aric.cscc.unc.edu/aric9/home | 1987-Present | Cohort | ARIC is one of the longest-running cohorts to understand the determinants of cardiovascular disease among US adults. Nearly 16,000 participants (45-65 years) from four communities (Washington MD, Forsyth NC, Jackson MS, Minneapolis, MN) were followed as part of the Main study and several Ancillary studies to characterize clinical and environmental risk factors. | In 2021, approximately 6,000 original participants are still active. |
| Coronary Artery Risk Development in Young Adults Study (CARDIA)  https://www.cardia.dopm.uab.edu/ | 1985-Present | Cohort | The CARDIA Study focuses on understanding the development and determinants of cardiovascular disease in young adults. It enrolled 5,115 black and white participants aged 18-30 at baseline, from four U.S. cities (Birmingham, AL; Chicago, IL; Minneapolis, MN; and Oakland, CA).(34) The mean BMI was 24.5 kg/m^2^ and the average fasting glucose (FPG) was 82 mg/dL at baseline. The study includes extensive cardiovascular assessments, lifestyle and behavioral surveys, and psychosocial measures. Follow-up examinations occur every 2-5 years. Significant findings have highlighted the impact of lifestyle factors such as diet, physical activity, and smoking on cardiovascular health, as well as the role of socioeconomic and psychosocial factors in disease risk. | After 25 years, CARDIA retained approximately 72% of surviving participants at the Year 25 examination and maintained >90% contact between exams and >80% contact at any two-year interval.(35) |
| Diabetes Prevention Program (DPP)  https://www.niddk.nih.gov/about-niddk/research-areas/diabetes/diabetes-prevention-program-dpp | 1996-2001 | RCT | The DPP aimed to determine whether lifestyle intervention or the medication metformin could prevent or delay the onset of type 2 diabetes in high-risk individuals. The study included 3,234 participants aged ≥25 years with elevated FPG or impaired glucose tolerance. The average age was 51 years, with about 68% women. The trial intentionally recruited a diverse cohort: 55% White (non-Hispanic), 20% Black, 16% Hispanic, 5% American Indian, and 4% Asian American.(36) The mean BMI was 34.0 kg/m², the average FPG was 106–108 mg/dL and mean HbA1c was 5.9%. Interventions focused on weight loss through diet and physical activity or metformin treatment. The study found that lifestyle intervention reduced diabetes incidence by 58%, and metformin by 31%, compared to placebo. Follow-ups assessed changes in diabetes status, metabolic measures, and cardiovascular risk factors. | During the core trial (1996–2002), retention was exceptionally high at about 94%, with >90% of participants completing assigned procedures and only ~2–3% lost due to death or withdrawal.(37) |
| Diabetes Prevention Program Outcomes Study (DPPOS)  https://dppos.bsc.gwu.edu/ | 2002 – present | RCT Follow-up of DPP | The DPPOS is a long-term follow-up of DPP participants to evaluate the sustained effects of the initial interventions on diabetes prevention and related health outcomes. It examines the long-term impact on diabetes incidence, cardiovascular outcomes, microvascular complications, and quality of life. The study continues to provide insights into the durability of lifestyle and pharmacologic interventions in managing and preventing type 2 diabetes over extended periods. | In long‐term follow‑up (20 years plus of combined DPP/DPPOS), around 84% of surviving participants continued annual visits (with 86–99% attendance at each) and >99% of planned data collected at those visits. |
| Jackson Heart Study (JHS)  https://www.jacksonheartstudy.org/ | 1998-Present | Cohort | The JHS is a large single-site, prospective epidemiologic investigation focusing on identifying genetic and environmental risk factors associated with the disproportionate burden of cardiovascular disease in African Americans. The study involves 5,301 participants aged between 21 and 94 years from Jackson, Mississippi, metropolitan area of Hinds, Madison, and Rankin Counties.(38) The initial baseline examination took place from 2000 to 2004, followed by two subsequent examinations from 2005 to 2008 and from 2009 to 2012. Follow-up examinations are conducted every three to four years, with ongoing annual follow-up interviews and cohort surveillance to track health outcomes and changes in risk factors. | JHS recruited and sought to retain about 6,500 African‑American adults via culturally tailored, community-based outreach, though publicly reported retention rates by exam cycle are not available. |
| Multi-Ethnic Study of Atherosclerosis (MESA)  https://www.mesa-nhlbi.org/ | 2000-Present | Cohort | MESA investigates the prevalence, causes, and progression of subclinical cardiovascular disease in a diverse, multi-ethnic population. The study includes 6,814 participants aged between 45 and 84 years from six U.S. communities (Los Angeles, CA; St. Paul, MN; New York, NY; Baltimore, MD; Chicago, IL; and Winston-Salem, NC), with no clinical cardiovascular disease at baseline. The cohort consisted of 53% female and 38.5% White, 27.8% African American, 21.9% Hispanic, and 11.8% Chinese American.(39) The mean age at baseline was around 62 years. The mean BMI at baseline was about 28 kg/m². Assessments involve extensive imaging studies (e.g., CT scans, MRI) and biomarker measurements. Follow-up exams every two to four years monitor the development of cardiovascular disease and risk factors. The study has contributed to understanding ethnic differences in cardiovascular risk and the role of subclinical disease in predicting clinical events. | MESA enrolled ~6,814 participants aged 45–84 without CVD, achieving ~96% enrollment of eligible individuals, and retained ~92% attendance at scheduled exams in the first decade—longer-term retention beyond year 10 is less consistently reported.(40) |

### Supplementary Table 2. Cohort data collection timeline

| **Cohort** | **Visit** | **Calendar year** | **Description (measurements taken, how many people are involved, regular follow-up or special event)** |
| --- | --- | --- | --- |
| ARIC | V1 | 1987–1989 | 15,792 participants; baseline exam; measurements: blood pressure, anthropometry, fasting blood/serum labs, ECG, medical history & medications, diet and physical-activity questionnaires. |
|  | V2 | 1990-1992 | 14,348 participants; 3-year follow-up exam; measurements: blood pressure, anthropometry, fasting blood/serum labs, ECG, medical history & medications, diet and physical-activity questionnaires. |
|  | V3 | 1993-1995 | 12,887 participants; 6-year follow-up exam; measurements: blood pressure, anthropometry, fasting blood/serum labs, ECG, medical history & medications, diet and physical-activity questionnaires |
|  | V4 | 1996-1998 | 11,656 participants; 9-year follow-up exam; measurements: blood pressure, anthropometry, fasting blood/serum labs, ECG, medical history & medications, diet and physical-activity questionnaires |
|  | V5 | 2011-2013 | 6538 participants; late-life exam; measurements: blood pressure, anthropometry, fasting blood/serum labs, ECG, medical history & medications, diet and physical-activity questionnaires; echocardiography and comprehensive cognitive testing |
|  | V6 | 2016-2017 | 4217 participants; late-life exam; measurements: blood pressure, anthropometry, fasting blood/serum labs, ECG, medical history & medications, diet and physical-activity questionnaires; echocardiography and comprehensive cognitive testing |
| CARDIA | Y0 | 1985-1986 | 5115 observations; baseline exam; measurements: blood pressure, laboratory measurements (plasma, serum), anthropometry, medical history, sociodemographics, family history, physical activity/fitness, diet questionnaires, psychological questionnaires, pulmonary function |
|  | Y2 | 1987-1988 | 4624 observations; year 2 follow-up; measurements: blood pressure, laboratory measurements (plasma, serum), anthropometry, medical history, sociodemographics, physical activity/fitness, diet questionnaires, psychological questionnaires, pulmonary function |
|  | Y5 | 1990-1991 | 4352 observations; year 5 follow-up; measurements: blood pressure, laboratory measurements (genetics, plasma, serum), anthropometry, medical history, sociodemographics, physical activity/fitness, obesity/eating questionnaires, psychological questionnaires, pulmonary function, echocardiography |
|  | Y7 | 1992-1993 | 4086 observations; year 7 follow-up; measurements: blood pressure, laboratory measurements (plasma, serum), anthropometry, medical history, sociodemographics, family history, physical activity/fitness, diet questionnaires, obesity/eating questionnaires, psychological questionnaires |
|  | Y10 | 1995-1996 | 3950 observations; year 10 follow-up; measurements: blood pressure, laboratory measurements (genetics, plasma, serum, urine), anthropometry, medical history, sociodemographics, family history, physical activity/fitness, diet questionnaires, obesity/eating questionnaires, psychological questionnaires, pulmonary function |
|  | Y15 | 2000-2001 | 3672 observations; year 15 follow-up; measurements: blood pressure, laboratory measurements (genetics, plasma, serum, urine), anthropometry, medical history, sociodemographics, physical activity/fitness, diet questionnaires, psychological questionnaires, pulmonary function, coronary calcium |
|  | Y20 | 2005-2006 | 3549 observations; year 20 follow-up; measurements: blood pressure, laboratory measurements (plasma, serum), anthropometry, medical history, sociodemographics, family history, physical activity/fitness, diet questionnaires, psychological questionnaires, pulmonary function |
|  | Y25 | 2010-2011 | 3599 observations; year 25 follow-up; measurements: blood pressure, laboratory measurements (plasma, serum), anthropometry, medical history, sociodemographics, physical activity/fitness, diet questionnaires, psychological questionnaires, pulmonary function |
|  | Y30 | 2015-2016 | 3358 observations; year 30 follow-up; measurements: blood pressure, laboratory measurements (genetics, plasma, serum), anthropometry, medical history, sociodemographics, physical activity/fitness, obesity/eating questionnaires, psychological questionnaires, pulmonary function, echocardiography |
| DPP | Baseline | 1994-2002 | 3,234 observations; participants randomized at 27 U.S. centers (Lifestyle = 1,079; Metformin = 1,073; Placebo = 1,082); measurements: blood pressure, anthropometry, fasting blood/serum labs including OGTT, ECG as indicated, medical history & medications, and diet/physical-activity questionnaires |
| DPPOS | Bridge | 2002-2003 | 2,766 observations; bridge follow-up to DPP; patients enrolled into DPPOS (~88% of eligible DPP participants); clinic visit to re-establish follow-up; measurements: blood pressure, anthropometry, blood/serum tests (glucose-related labs), ECG where indicated, medical history & medications, and lifestyle questionnaires |
|  | Phase 1 | 2002-2008 | 2,766 observations; participants enrolled into DPPOS; Participants from Bridge attended annual follow-ups; measurements: blood pressure, anthropometry, blood/serum tests focused on glycemia, ECG where indicated, medical history & medications, and lifestyle/quality-of-life questionnaires |
|  | Phase 2 | 2008-2015 | **Continuation of DPPOS cohort (analysis counts vary by year) with added emphasis on microvascular outcomes (retinopathy, nephropathy, neuropathy) and atherosclerosis imaging** |
|  | Phase 3 | 2016-2020 | **Continuation of DPPOS cohort (analysis counts vary by year) with added emphasis on macrovascular complications, cancer, and mortality** |
| JHS | V1 | 2000-2004 | 5,306 observations; baseline exam; measurements: blood pressure, anthropometry, fasting blood/serum labs, ECG, medical history & medications, diet and physical-activity questionnaires. |
|  | V2 | 2005-2008 | 4,203 observations; 5-year follow-up; measurements: blood pressure, anthropometry, fasting blood/serum labs, ECG, medical history & medications, diet and physical-activity questionnaires. |
|  | V3 | 2009-2013 | 3,819 observations; 4-year follow-up; measurements: blood pressure, anthropometry, fasting blood/serum labs, ECG, medical history & medications, diet and physical-activity questionnaires. |
| MESA | V1 | 2000-2002 | 6814 observations; baseline exam; measurements: anthropometry, diet, echocardiography, health and life, medical history, medications, neighborhood, personal history, physical activity, seated blood pressure |
|  | V2 | 2002-2004 | 6232 observations; 2-year follow up; measurements: anthropometry, eye history, family history, health and life, medical history, medications, personal history, physical activity, seated blood pressure, sleep history |
|  | V3 | 2004-2005 | 5939 observations; 4-year follow up; measurements: anthropometry, health and life, medical history, medications, personal history, physical activity, seated blood pressure |
|  | V4 | 2005-2007 | 5704 observations; 6-year follow up; measurements: anthropometry, health and life, medical history, medications, personal history, seated blood pressure, sleep history |
|  | V5 | 2010-2011 | 4655 observations; 10-year follow up; anthropometry, health and life, medical history, medications, personal history, seated blood pressure, physical activity, food frequency diet, erectile dysfunction, cognitive assessment, digit span, digit symbol |

### Supplementary Table 3. Measurement protocol of key variables

| **Cohort** | **Measure** | **Protocol** |
| --- | --- | --- |
| ARIC | Height and Weight | Standing height and weight are measured at enrollment and most study visits. Scale to measure body weight used in Detecto Model #437. Metal anthropometric rulers in centimeters (200cm) used to measure height. |
| CARDIA | Height and Weight | Height (anthropometric ruler in centimeters or Stadiometer) and weight (Detecto Scale Model#68965) were measured at baseline and all study follow-up visits. |
| DPP | Height and Weight | Both height and weight were measured at the baseline and end of study. Weight was measured every 6 months. |
| DPPOS | Height and Weight | Weight was measured at 6-month intervals. Height was measured at the first annual visit and every 5-6 years. |
| JHS | Height and Weight | Weight was measured by a balance scale (Detector, model #437) and height was measured by traditional wall mounted tape at baseline and follow-up study visits. Tanita TBF 300A Body Composition Analyzer scale was used to assess the comparability of measurements. |
| MESA | Height and Weight | Weight was measured using a Detecto Platform Balance Scale and height was measured with stadiometer or height ruler at each study visit. |
| ARIC | HbA_1c_ (%) | HbA_1c_ was measured using high-performance liquid chromatography (HPLC) methods, standardized to the Diabetes Control and Complications Trial assay. Visit 5 was the first visit at which HbA_1c_ was measured. |
| CARDIA | HbA_1c_ (%) | HbA_1c_ was measured at some study exams. |
| DPP | HbA_1c_ (%) | HbA_1c_ was measured at baseline, end of study, and 6 or 12-month intervals. |
| DPPOS | HbA_1c_ (%) | HbA_1c_ was measured at annual visits. |
| JHS | HbA_1c_ (%) | HbA_1c_ was measured using a National Glycohemoglobin Standardization Program (NGSP) certified assay at study baseline visit and follow up visits. |
| MESA | HbA_1c_ (%) | Fasting lipid profile was measured at each study visit. HDL cholesterol was measured using the cholesterol oxidase method (Roche Diagnostics, Indianapolis, IN, USA). Triglyceride concentrations were measured using a glycerol-blanked enzymatic method LDL cholesterol was calculated using the Friedewald formula. |
| ARIC | Fasting glucose | The Central Clinical Chemistry Laboratory at the University of Minnesota processed the frozen blood serum samples collected from field centers. Participants were asked to fast prior to venipuncture. Fasting glucose was measured at enrollment and some study visits. |
| CARDIA | Fasting glucose | Fasting glucose was measured by hexokinase ultraviolet method at some study exams. |
| DPP | Fasting glucose | A blood sample (after 12 hours of fasting) was taken from the arm to measure fasting glucose levels every 6 months. |
| DPPOS | Fasting glucose | Fasting glucose was measured at both mid-year and annual visits. |
| JHS | Fasting glucose | Fasting glucose (8 hours of fasting) was measured by glucose oxidase method as duplicates in the baseline study visit and follow up visits. |
| MESA | Fasting glucose | Blood sample drawn at fasted state at all study visits. |
| ARIC | Fasting insulin | The Central Clinical Chemistry Laboratory at the University of Minnesota processed the frozen blood serum samples collected from field centers. Participants were asked to fast prior to venipuncture. Fasting insulin was measured at enrollment and some study visits. |
| CARDIA | Fasting insulin | Fasting insulin was measured by radioimmunoassay at some study exams. |
| DPP | Fasting insulin | Fasting insulin and fasting plasma proinsulin were collected at baseline, end of study, and every 12 months. |
| DPPOS | Fasting insulin | Fasting insulin was measured at mid-year visits. |
| JHS | Fasting insulin | Fasting plasma insulin were measured by total immunoreactive assay and standardized to serum levels at baseline study visits and follow up visits. |
| MESA | Fasting insulin | Blood sample drawn at fasted state at all study visits. |
| ARIC | Lipid panel | Lipid measurements were performed by the Central Lipid Laboratory. At baseline and multiple study visits, plasma cholesterol, triglycerides, HDL cholesterol and LDL cholesterol, plasma ApoA-I and apoB were collected. |
| CARDIA | Lipid panel | Lipid assays were conducted at each study exam including total cholesterol by trinder-type method and determined enzymatically on the Abbot Spectrum (using Hitachi 917 – R1 cholesterol reagent); HDL cholesterol by trinder-type method and determined enzymatically after dextran sulfate –magnesium precipitation on the Abbot Spectrum; and, triglycerides by ultraviolet method and determined enzymatically on the Abbot Spectrum (using Hitachi 917 – R1Buffer/4- Chloropheno/Enzymes). LDL cholesterol was calculated using the Friedewald equation. |
| DPP | Lipid panel | Fasting lipid profile (total cholesterol, total triglyceride, HDL-cholesterol and derived LDL-cholesterol) was measured at baseline, end of study, and at 6 or 12-month intervals. In cases of hypertriglyceridemia, beta quantification specifically measuring LDL-cholesterol is performed. |
| DPPOS | Lipid panel | Fasting lipid profile (total cholesterol, total triglyceride, HDL-cholesterol and derived LDL-cholesterol) was measured annually. In cases of hypertriglyceridemia, beta quantification specifically measuring LDL-cholesterol is performed. |
| JHS | Lipid panel | Routine plasma lipid tests (cholesterol, triglycerides, and HDL-cholesterol) were conducted at each study visit. |
| MESA | Lipid panel | Blood lipids (total cholesterol, HDL and LDL cholesterol, and triglycerides) and lipoproteins were measured at the Central Lipid Laboratory. |
| ARIC | Blood Pressure | The sitting arm blood pressure is measured three times at each clinic visit. It takes about 10-15 minutes to make three blood pressure measurements including the initial five minutes rest. |
| CARDIA | Blood Pressure | Seated BP is measured three times at each clinic visit. The seated BP reading is the average of the second and third systolic and diastolic BPs calculated by computer. |
| DPP | Blood Pressure | Blood pressure was measured in baseline, end of study, and in arm every 6 months and in ankle every 12 months. |
| DPPOS | Blood Pressure | Blood pressure was measured in arm at mid-year and annual visits and in ankle annually. |
| JHS | Blood Pressure | Sitting blood pressure were measured in a resting state, using 2 measurements with a random zero sphygmomanometer at each study visit. |
| MESA | Blood Pressure | Resting blood pressure was measured in the right arm  in the seated position by an automated oscillometric method (Dinamap) at all study exams. Three readings were taken; the second and third readings were averaged to obtain the blood pressure levels used in analyses. |
| ARIC | Serum creatinine | Creatinine was measured using the DART Creatinine reagent based on a modified kinetic Jaffe method, in which creatinine reacts with picrate in an alkaline solution to form a red creatinine–picrate complex. Absorbance was measured at 520 nm and quantified using an initial-rate algorithm with regression-based calibration against a reagent blank. Outlier rejection procedures were applied; no post-processing corrections were performed. |
| CARDIA | Serum creatinine | Creatinine was measured by a modified-rate Jaffe  method and assayed by kinetic in vitro tests using rate-blanking and compensation for the quantitative determination of creatinine in human serum and  plasma; kinetic, substrate triggered, rate-blanked method |
| DPP | Serum creatinine | Yearly serum creatinine measurements were performed in the MET and PLB groups for safety, allowing for calculation of estimated GFR (eGFR). All the laboratory measurements were performed at the Central Biochemistry Laboratory (Northwest Lipid Research Laboratories, University of Washington, Seattle, WA). Pertinent to the current analyses, creatinine concentrations in the serum and urine were measured by a variation of the Jaffe method traceable to isotope-dilution mass spectrometry |
| DPPOS | Serum creatinine | During DPPOS, ACR and serum creatinine measurements were performed yearly in all three treatment groups. All the laboratory measurements were performed at the Central Biochemistry Laboratory (Northwest Lipid Research Laboratories, University of Washington, Seattle, WA). Pertinent to the current analyses, creatinine concentrations in the serum and urine were measured by a variation of the Jaffe method traceable to isotope-dilution mass spectrometry |
| JHS | Serum creatinine | Creatinine was measured with a multipoint enzymatic spectrophotometric assay at the baseline visit (2000–2004).(41) The calibration equation was applied to creatinine measurements of 5,210 participants to estimate glomerular filtration rate |
| MESA | Serum creatinine | Creatinine was measured by rate reflectance spectrophotometry using thin-film adaptation of the creatinine amidinohydrolase method on the Vitros analyzer (Johnson & Johnson Clinical Diagnostics Inc., Raritan, NJ) at the Collaborative Studies Clinical Laboratory at Fairview-University Medical Center (Minneapolis, MN) and calibrated to the Cleveland Clinic. (42) In REGARDS, creatinine was also measured by a Vitros analyzer and calibrated to an international isotope dilution mass spectroscopic-traceable standard |
| ARIC | Diabetes definition | Fasting blood glucose ≥126 mg/dl (7.0 mmol/L), non-fasting blood glucose ≥200 mg/dl (11.1 mmol/L), self-reported physician diagnosis of diabetes, or self-reported use of diabetes medications. |
| CARDIA | Diabetes definition | Fasting blood glucose ≥126 mg/dl (7.0 mmol/L), non-fasting blood glucose ≥200 mg/dl (11.1 mmol/L), self-reported physician diagnosis of diabetes, or self-reported use of diabetes medications. |
| DPP | Diabetes definition | Fasting plasma glucose level ≥126 mg/dL (7.0 mmol/L), or 75-gram OGTT resulting in 2-hour plasma glucose ≥ 200 mg/dL (11.1 mmol/L). |
| DPPOS | Diabetes definition | Fasting plasma glucose level ≥ 126 mg/dL [7.0 mmol/L] or 2-hour plasma glucose ≥ 200 mg/dL [11.1 mmol/L], after a 75-gram OGTT, and confirmed with a repeat test. |
| JHS | Diabetes definition | Current use of insulin or oral antidiabetic agent, OR self-report of physician’s diagnosis or fasting glucose ≥ 126 mg/dl, or hemoglobin A1c ≥ 6.5%. |
| MESA | Diabetes definition | Fasting blood glucose ≥126 mg/dl (7.0 mmol/L) OR participants self-reporting a physician-diagnosed history of diabetes and the use of insulin or oral hypoglycemic medications. |
| ARIC | ASCVD Definition | ASCVD was defined as the first occurrence of coronary heart disease (CHD), myocardial infarction (MI), or stroke. Events were ascertained from main study visits (v1–v7) and the cohort annual follow-up (AFU) datasets. For each participant, the ASCVD event date was the earliest visit/AFU record at which an ASCVD event was recorded; participants without any recorded ASCVD event were censored at their last available visit/AFU record. |
| CARDIA | ASCVD Definition | ASCVD was defined as the first occurrence of coronary heart disease (CHD), myocardial infarction (MI), or stroke. ASCVD events were ascertained from the study outcomes dataset, which prospectively captures adjudicated cardiovascular events occurring between study examinations (and after baseline) during follow-up. For each participant, the ASCVD event date was the earliest outcomes record indicating an ASCVD event; participants without any recorded ASCVD event were censored at their last available outcomes record. |
| DPP | ASCVD Definition | ASCVD was defined as the first occurrence of myocardial infarction (MI), or stroke. In DPP, ASCVD history and incident events were identified from the screening dataset and the follow-up questionnaires dataset, which capture participant-reported (and study-recorded) cardiovascular diagnoses during study follow-up. For each participant, the ASCVD event date was assigned as the earliest record in either dataset indicating an ASCVD event; participants with no recorded ASCVD event were censored at their last available screening/questionnaire record. |
| DPPOS | ASCVD Definition | ASCVD was defined as incident myocardial infarction (MI) only. MI events were ascertained from the DPPOS_Phase1 datasets, and for each participant the MI event date was assigned as the earliest record indicating an MI; participants without any recorded MI were censored at their last available DPPOS_Phase1 record. |
| JHS | ASCVD Definition | ASCVD was defined as the first occurrence of coronary heart disease (CHD), myocardial infarction (MI), or stroke. Events were ascertained from main study visits (v1–v3) and the cohort annual follow-up (AFU) datasets. For each participant, the ASCVD event date was the earliest visit/AFU record at which an ASCVD event was recorded; participants without any recorded ASCVD event were censored at their last available visit/AFU record. |
| MESA | ASCVD Definition | ASCVD was defined as the first occurrence of coronary heart disease (CHD), myocardial infarction (MI), or stroke. Events were ascertained from the MESA cardiovascular (CVD) event dataset, which prospectively records adjudicated CVD outcomes occurring during longitudinal follow-up, including events identified between and after in-person study visits. For each participant, the ASCVD event date was assigned as the earliest CVD event record indicating CHD, MI, or stroke; participants without any recorded ASCVD event were censored at their last available CVD event record. |

### Supplementary Table 4. Descriptive characteristics by cluster in the validation sample

|  | **Overall** | ***Cluster A*** | ***Cluster B*** |
| --- | --- | --- | --- |
| *N* | 3250 | 2677 | 570 |
| **First visit** |  |  |  |
| Age | 54.9 (6) | 55.1 (6) | 51.3 (4.9) |
| Female (%) | 1751 (53.9%) | 1619 (53.1%) | 132 (65%) |
| Race (%) ^a^ |  |  |  |
| *Black* | 817 (25.1%) | 771 (25.3%) | 46 (22.7%) |
| *White* | 1342 (41.3%) | 1222 (40.1%) | 120 (59.1%) |
| *Hispanic* | 695 (21.4%) | 680 (22.3%) | 15 (7.4%) |
| *Other* | 396 (12.2%) | 374 (12.3%) | 22 (10.8%) |
| Smoking status (%) |  |  |  |
| *Never or Unknown* | 1585 (48.8%) | 1495 (49.1%) | 90 (44.3%) |
| *Former* | 1131 (34.8%) | 1053 (34.6%) | 78 (38.4%) |
| *Current* | 534 (16.4%) | 499 (16.4%) | 35 (17.2%) |
| Alcohol use status (%) |  |  |  |
| *Never or Unknown* | 583 (17.9%) | 557 (18.3%) | 26 (12.8%) |
| *Former* | 235 (7.2%) | 229 (7.5%) | 6 (3%) |
| *Current* | 2432 (74.8%) | 2261 (74.2%) | 171 (84.2%) |
| Medication use |  |  |  |
| *Antihypertensive* | 788 (24.2%) | 779 (25.6%) | 9 (4.4%) |
| *Lipid lowering* | 346 (10.6%) | 336 (11%) | 10 (4.9%) |
| DPP Treatment Status |  |  |  |
| *Placebo/Non-DPP* | 3250 (100%) | 3047 (100%) | 203 (100%) |
| *Lifestyle* | - |  |  |
| *Metformin* | - |  |  |
| Key Biomarkers |  |  |  |
| *Body mass index (kg/m^2^)* | 28.2 (5.4) | 24.7 (3.8) | 28.2 (5.4) |
| *Systolic BP (mmHg)* | 120.7 (18.6) | 107.6 (12.8) | 120.7 (18.6) |
| *Diastolic BP (mmHg)* | 72.6 (10.3) | 67.1 (9.2) | 72.6 (10.3) |
| *Waist circumference (cm)* | 96.3 (14.3) | 84.9 (12.1) | 96.3 (14.3) |
| *LDL cholesterol (mg/dL)* | 119.4 (31.1) | 116.1 (29.1) | 119.4 (31.1) |
| *HDL cholesterol (mg/dL)* | 51.1 (14.8) | 59.4 (15.7) | 51.1 (14.8) |
| *Triglycerides (mg/dL)* | 128.5 (81) | 87.8 (38) | 128.5 (81) |
| *Total cholesterol (mg/dL)* | 196.1 (35.2) | 193.1 (31.7) | 196.1 (35.2) |
| *Triglyceride-HDL ratio* | 2.2 (1.4, 3.6) | 1.5 (0.9, 2) | 2.2 (1.4, 3.6) |
| *Serum creatinine (mg/dL)* | 0.9 (0.2) | 0.9 (0.2) | 0.9 (0.2) |
| *eGFR (mL/min/1.73 m²)* | 85.4 (75, 96.6) | 86.2 (76.1, 92.5) | 85.4 (75, 96.6) |
| *Fasting Insulin (µIU/mL)* | 9.5 (5.4) | 6.5 (2.9) | 9.5 (5.4) |
| *Fasting Glucose (mg/dL)* | 87.5 (9.3) | 80.6 (7.4) | 87.5 (9.3) |
| *HbA1c (%)* | - | - | - |
| *HOMA2-%B* | 95 (78, 119.5) | 95.3 (77.8, 120.2) | 91.4 (79.6, 110.9) |
| *HOMA2-IR* | 0.9 (0.6, 1.3) | 0.9 (0.7, 1.3) | 0.6 (0.5, 0.8) |
| **First 10 years** |  |  |  |
| Observations | 13099 | 12252 | 847 |
| Number of years with visits | 4 (4, 5) | 4 (4, 5) | 4 (4, 5) |
| Newly diagnosed diabetes | 292 (9.0%) | 289 (9.5%) | 3 (1.5%) |
| **Overall** |  |  |  |
| Observations | 17655 | 16494 | 1161 |
| Number of visits with biomarkers measured | 6 (5, 6) | 6 (5, 6) | 6 (6, 6) |
| Newly diagnosed diabetes | 487 (28.7%) |  |  |
| Age at diabetes diagnosis | 65.1 (7.4) | 65.2 (7.4) | 59.8 (11.3) |
| Follow-up time with biomarkers | 15 (10, 16) | 15 (10, 16) | 16 (15, 16) |
| Follow-up time till death or censoring (years) | 19.4 (18.7, 19.8) | 19 (19, 20) | 20 (19, 20) |

### Supplementary Figure 1. Description of training window and outcome assessment period

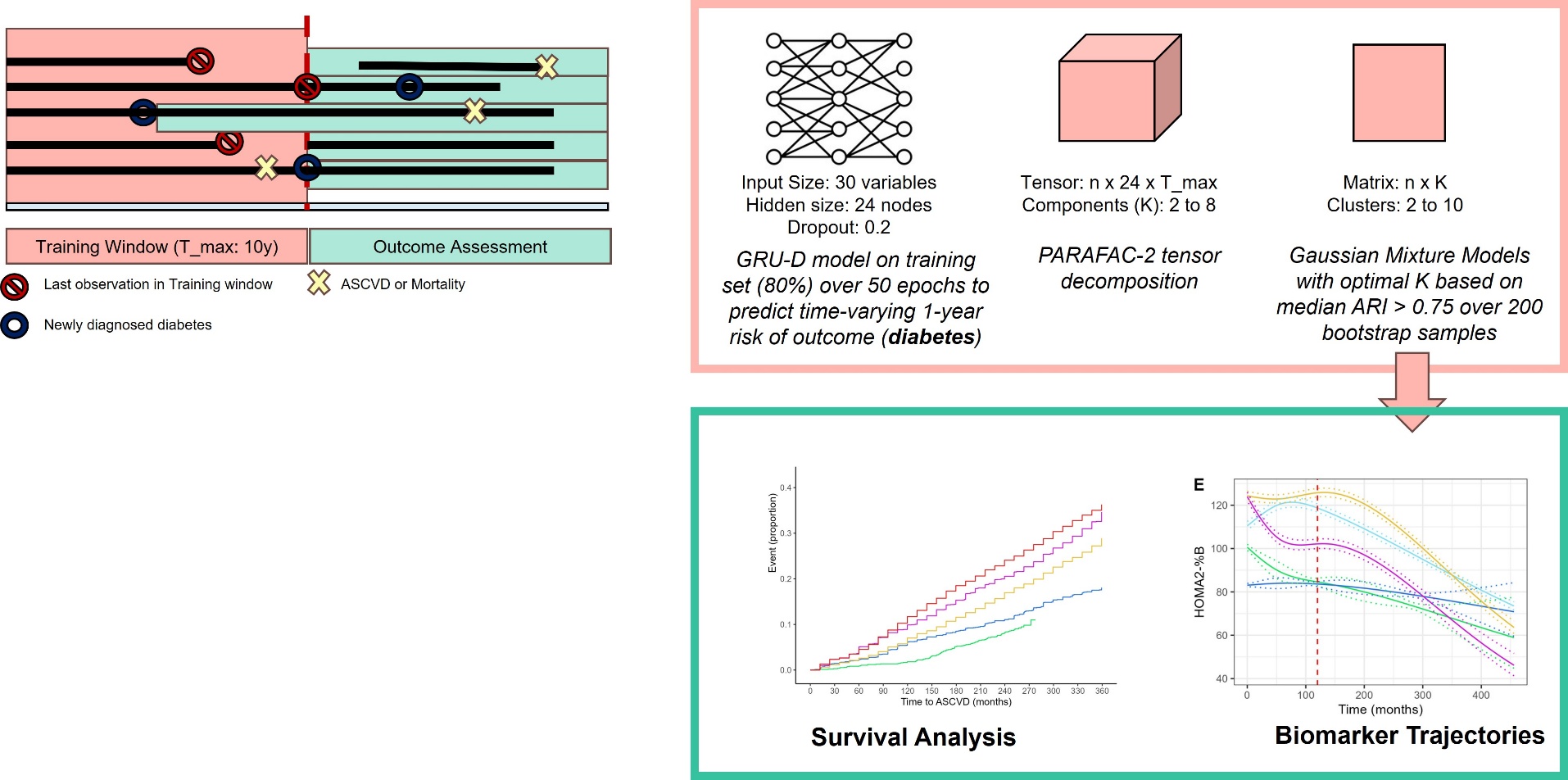

### Supplementary Figure 2. Flowchart of analytic sample

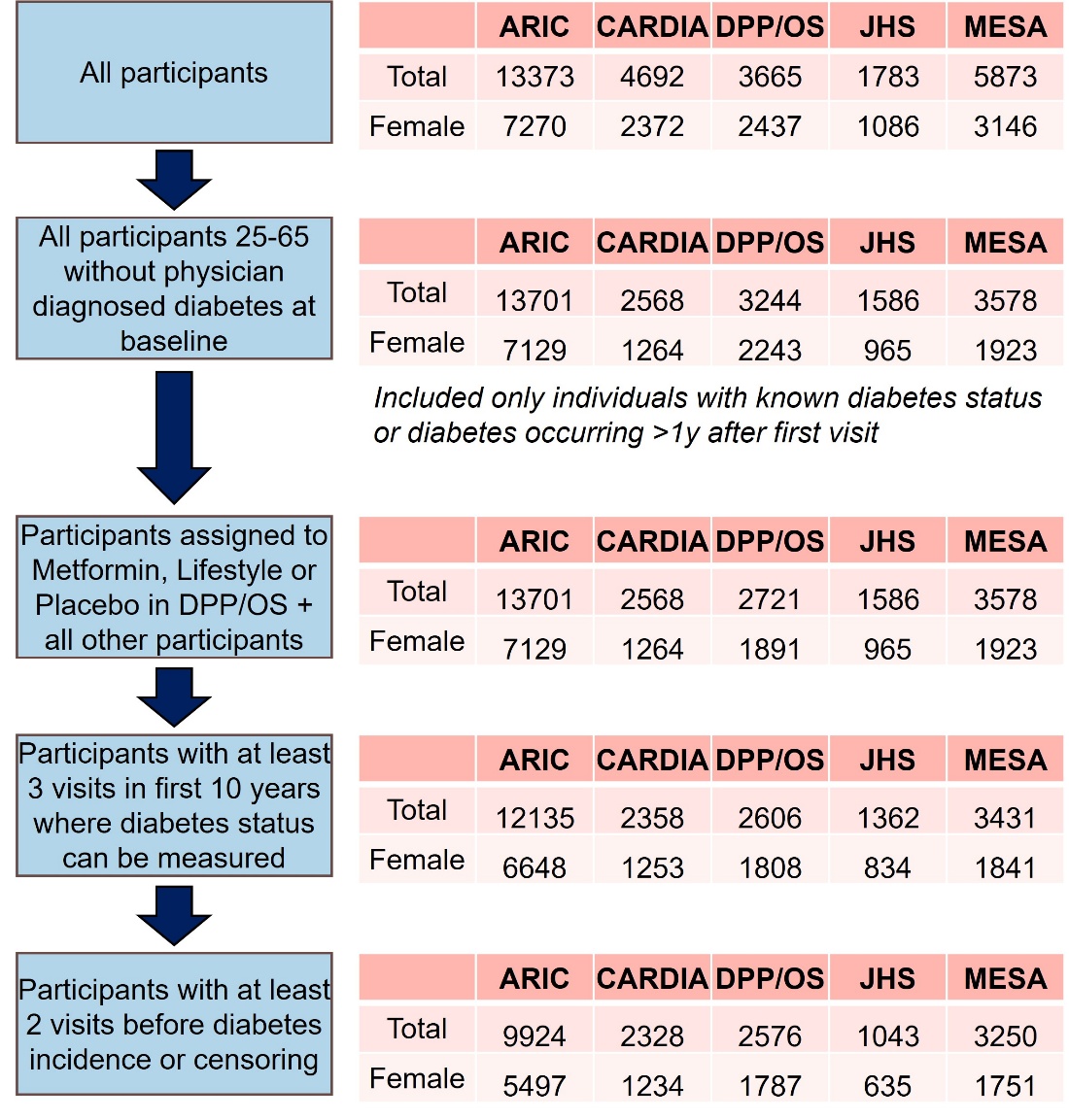

### Supplementary Figure 3. Time since baseline for study visits

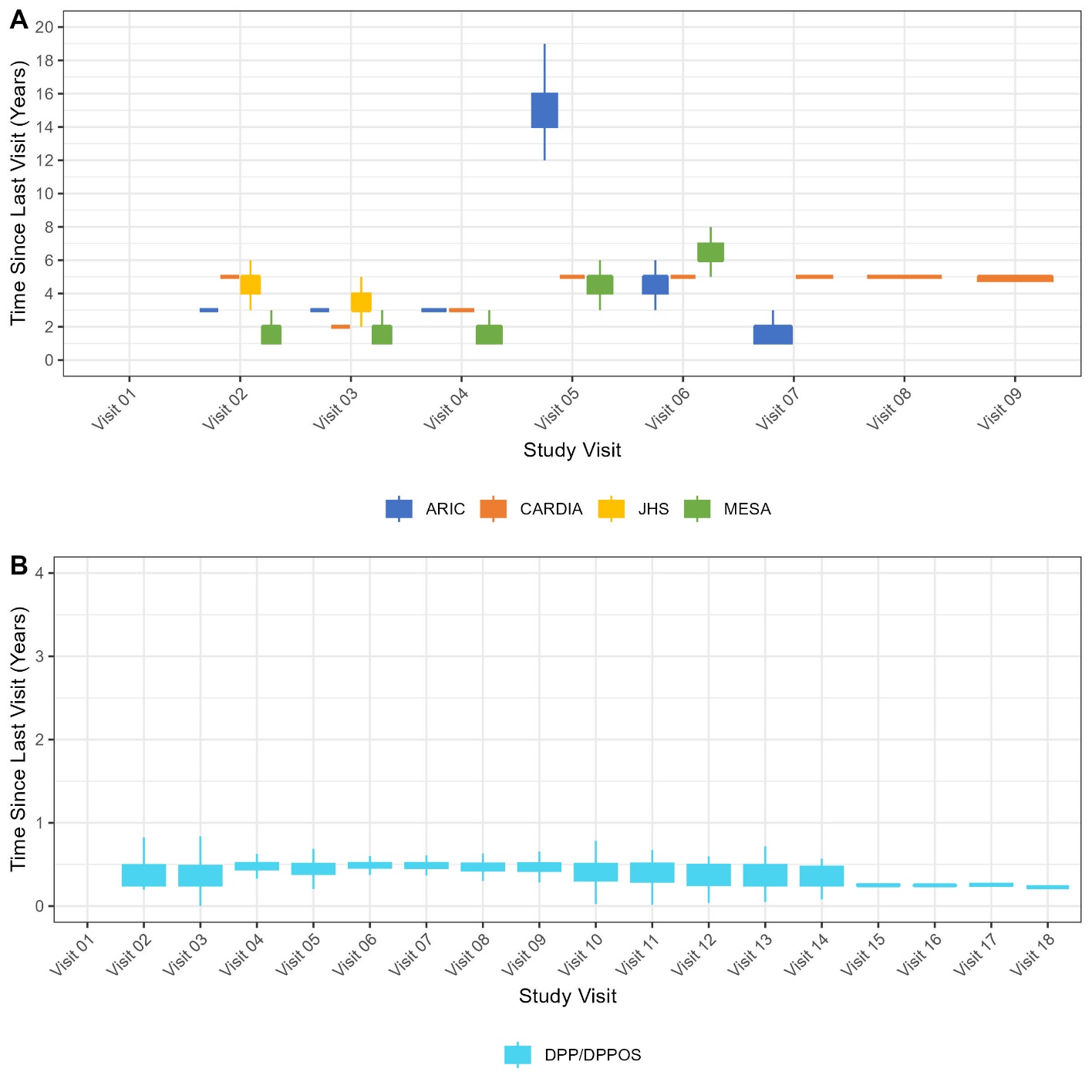

### Supplementary Figure 4. Cumulative time elapsed by visit and cohort

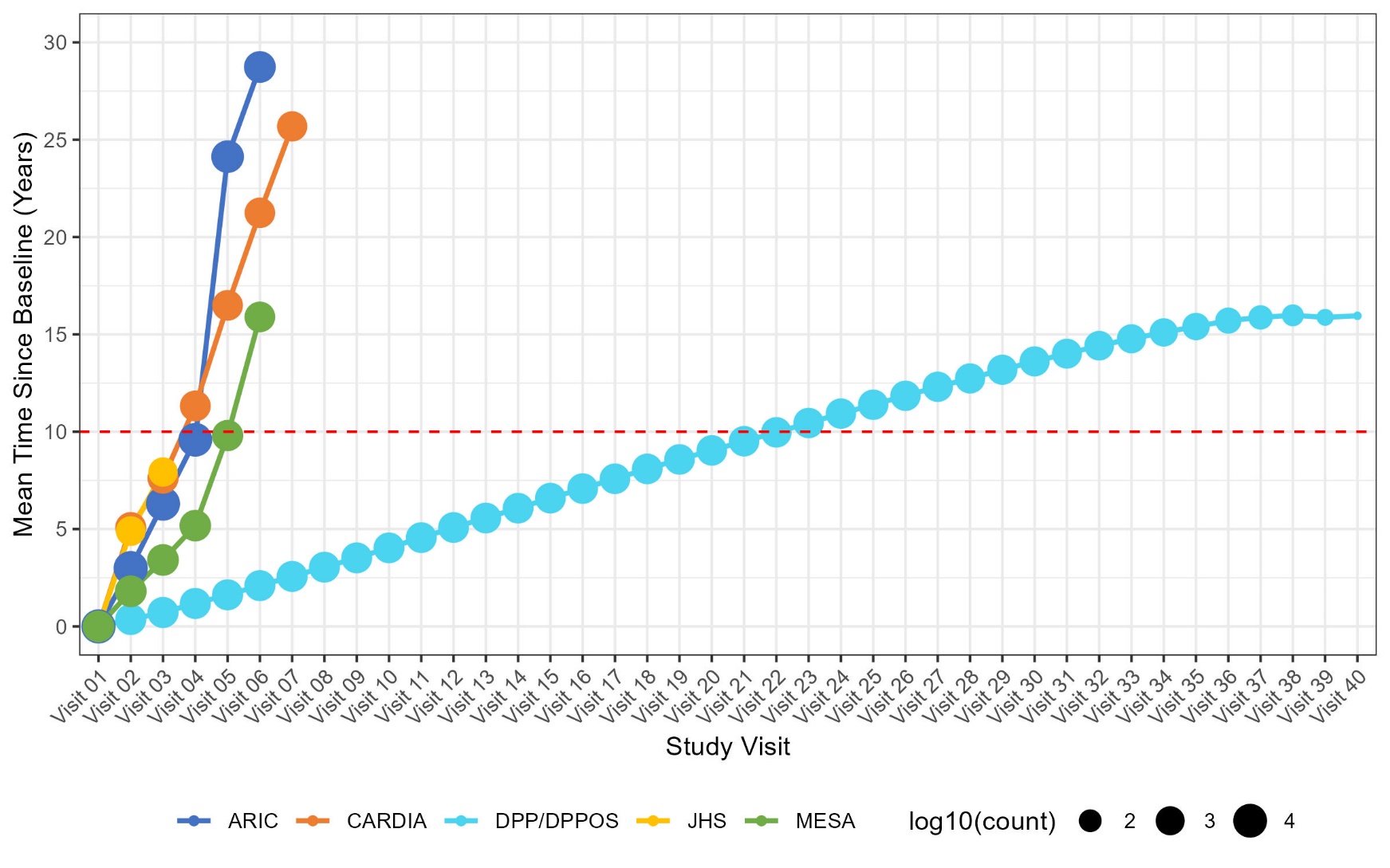

### Supplementary Figure 5. Data availability among analytic sample in first 10 years of follow-up

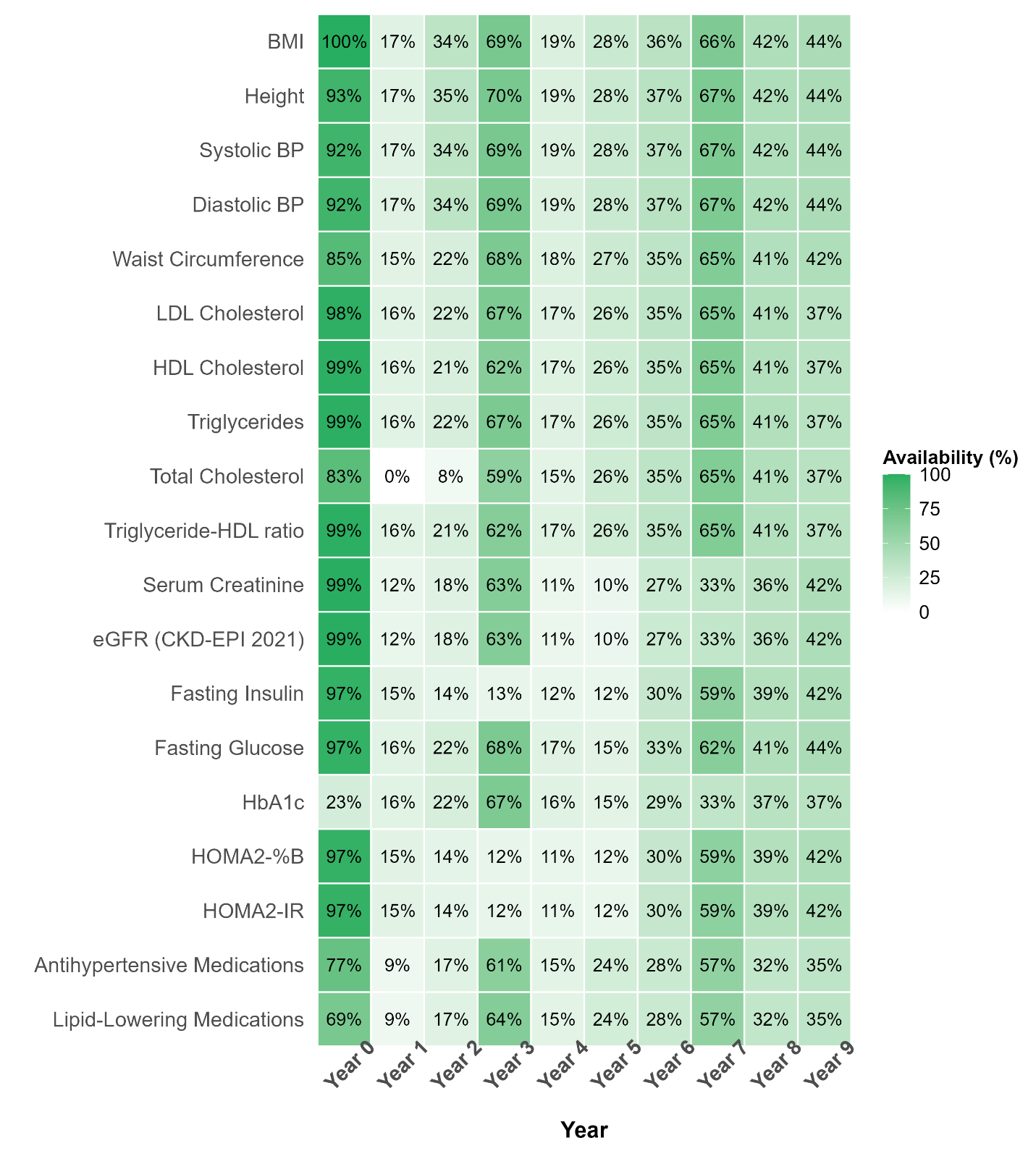

Information on how missingness is handled by GRU-D is provided in Supplementary Methods Note. Variables with extremely high rates of missingness across all observations, by virtue of the model framework do not contribute to prediction of diabetes risk.

### Supplementary Figure 6. Model performance in held-out training and validation datasets

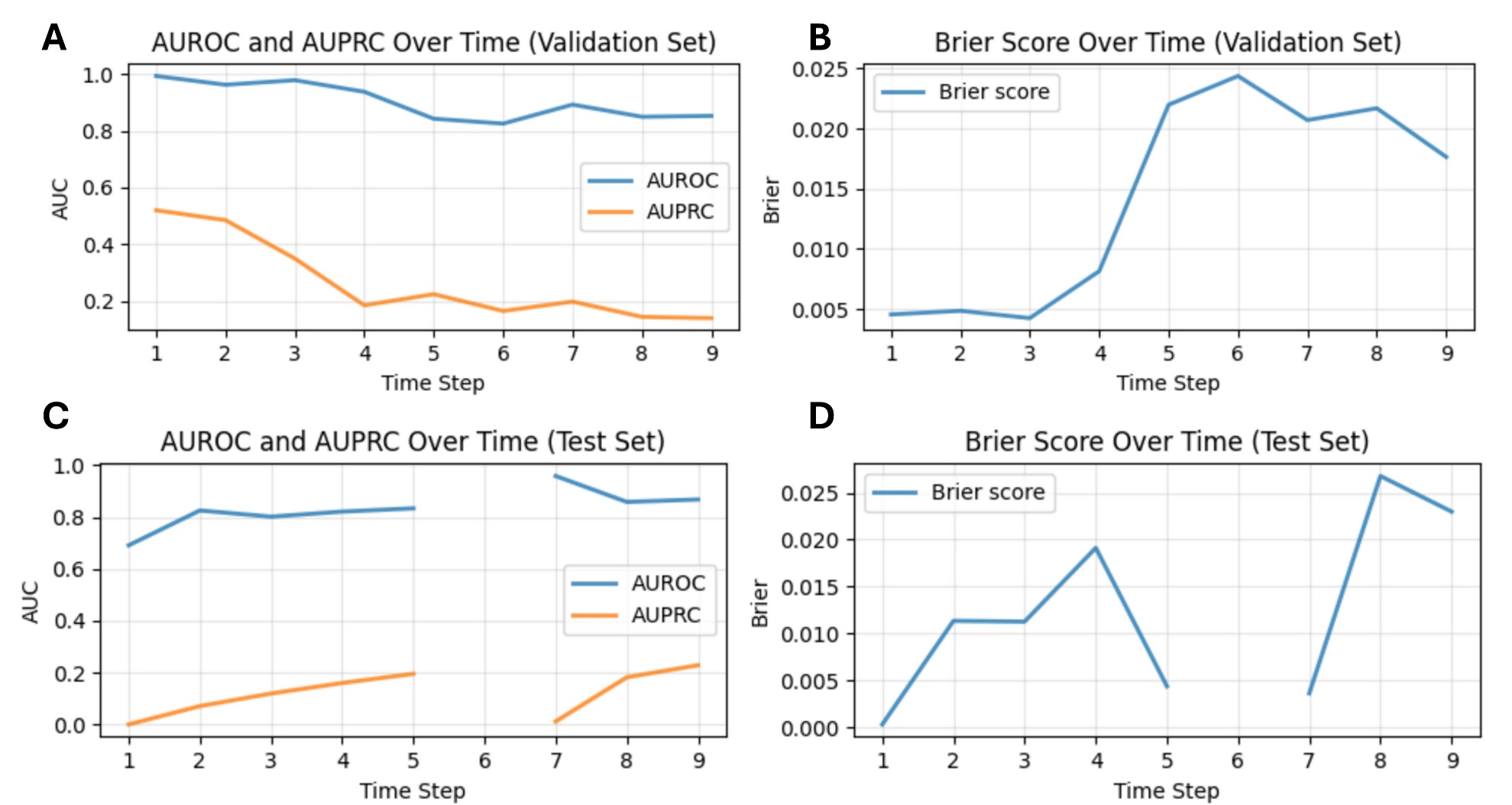

### Supplementary Figure 7. Identifying rank and number of clusters from neural network embeddings

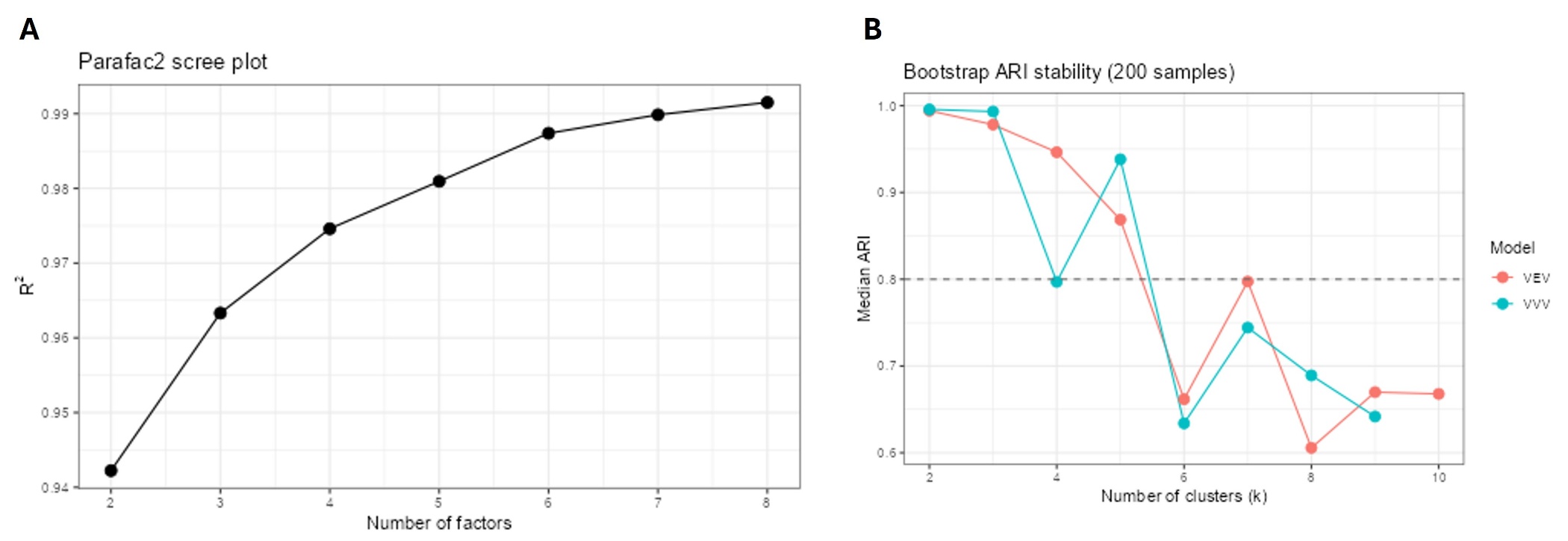

### Supplementary Figure 8. Distribution of socio-demographic and clinical characteristics at baseline by cluster

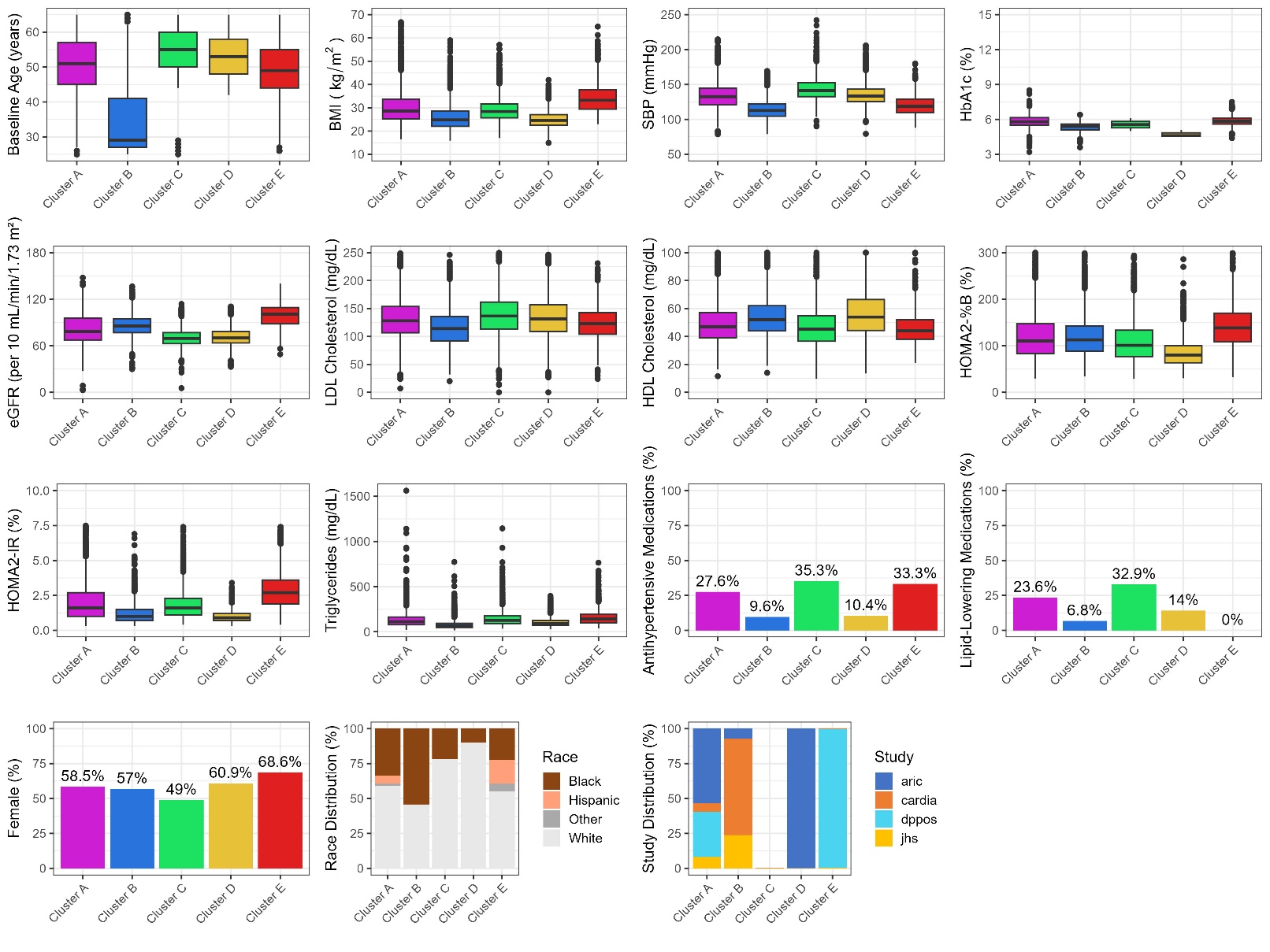

### Supplementary Methods Note

Variables (24 variables)

The following variables were time fixed across cohorts:

- *Binary*: Female sex
- *Categorical*: Race (Asian, Black, Hispanic, White, Other), Treatment (Metformin [DPP only], Lifestyle [DPP only], Placebo [Other])
- *Continuous*: Baseline Age

The following variables were longitudinally harmonized across pooled cohorts:

- *Binary*: Current medication use for high blood pressure, high cholesterol
- *Categorical*: Smoking use (never, former, current), Alcohol use (never, former, current)
- *Continuous*: BMI, Systolic BP, Diastolic BP, Waist circumference, LDL cholesterol, HDL cholesterol, Triglycerides, Total Cholesterol, Serum Creatinine, eGFR (CKD-EPI2021), Fasting insulin, Fasting glucose, HbA1c, HOMA2-%B, HOMA2-IR

Gated Recurrent Unit – Decay

We modeled time-to-event outcomes using a discrete-time hazard formulation, implemented via a Gated Recurrent Unit with Decay (GRU-D) to accommodate irregularly sampled longitudinal covariates and informative missingness. The approach is conceptually similar to a pooled logistic regression that allows us to estimate hazard ratios.

logit(Y_i,t_ = 1 | Y_i,t-1_ = 0, X_it-1_) = α_t-1_ + β_t_X_it-1_,

where α_t_ is the time-specific intercept or baseline hazard and X_it_ are the time-varying covariates.

Let ( i = 1, …, *N*) index individuals and ( t = 1, …, T_i_) index discrete follow-up intervals (e.g., years). Each individual contributes repeated measurements (with missing data) until the occurrence of the event (diabetes diagnosis) or censoring at T_max_..

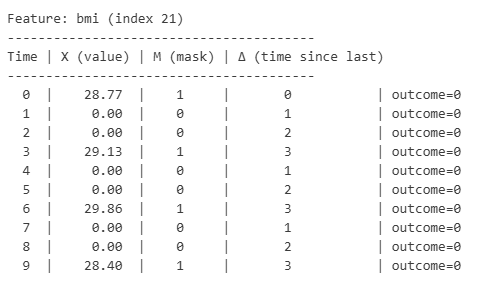

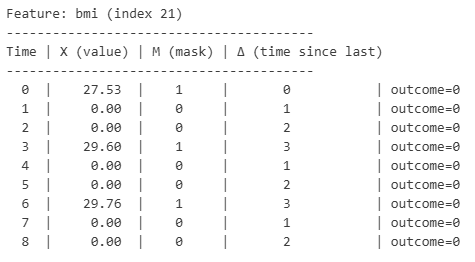

Sequences of BMI for two example patients are provided above.

The longitudinal data is transformed into a tensor of dimensions (*N, 3, D, T*) where N is the unique number of individuals, D is the unique number of variables, T is the maximum number of time steps (T_max_) and 3 indicates three datasets (X: raw input, M: masking vector, Δ: Time interval vector).

The learned parameters and their dimensions (**column, row**) are the following:

- W_γ.x_: Input decay rate for each of the input features **(D, D)**
- W_γ.h_: Hidden state decay rate for each of the units **(D, H)**
- W_r_: Reset gate weights to decide how much old information to retain **(2D + H, H)**
- W_z_: Update gate weights to decide how much new information to accept **(2D + H, H)**
- W_h_: Hidden state weights to compute new information **(2D + H, H)**
- W_out_: Mapping hidden state to output linear layer (i.e., logit scale) **(H, 1)**

Loss at time t is calculated using the Binary Cross-Entropy, after incorporating a mask for whether the participant is still at risk (i.e., no history of outcome and not censored). Each participant contributes equally to the loss regardless of their follow-up duration through a per-patient normalization step that only accounts for their ‘at risk’ time to handle right-censoring. This includes observations that are handled via time decay for those who were right censored at year 10.

- For input data: γ_xt_ = exp[-(max(0, W_γ.x_δt + b_γ_]
  - Measurement vector (x_decayed,dt_) for variable d and time t is then derived from observed data (x_dt_), masking vector (m_dt_), decay (γ_xt_) and the empirical mean ( $\tilde{x}$_dt_) – [Eq 11 in *Che 2018*]
- For hidden states: h_decayed_ = γ_h 🞊_ h_t-1_ – [Eq 12 in *Che 2018*]
  - γ_h_ is derived from W_γ.h_ and h_t-1_
- Standard GRU gates are applied to decayed inputs and hidden states
  - Reset gate – [Eq 13 in *Che 2018*]
  - Update gate – [Eq 14 in *Che 2018*]
  - Candidate hidden state – [Eq 15 in *Che 2018*]
  - Final hidden state – [Eq 16 in *Che 2018*]
- Logit output: W_out_.h_t_

The final model was trained using random dropout of 20% of hidden units during training for regularization to prevent overfitting using the Adaptive gradient method (Adam) optimizer, learning rate of 0.001, and weight decay of 10^-5^.

Parallel Factor Analysis-2 (PARAFAC-2)

Tensor decomposition of the longitudinal embeddings (n x H x T_max) was carried out using alternating least squares. PARAFAC-2, for a three-dimensional tensor, decomposes the input into three matrices: A (individuals x components x embeddings), B (time x components), C (embeddings x components).

We identified the optimal number of components (**R**) from the gradient of the R-squared, and was chosen to be six.

We extracted the first mode of the A tensor (n x R) for clustering. The first mode represents a low-rank representation of the heterogeneity of longitudinal 1-year diabetes risk profiles of individuals.

Gaussian Mixture Models

Gaussian Mixture Models (GMM) were used to cluster the components from the low-rank representation obtained from PARAFAC-2. GMMs model the probability of membership in cluster *k* (π*_k_*) with mean μ_k_ and covariance Σ_k_. We fit two sets of GMMs (VEV: Ellipsoidal, equal shape, VVV: Ellipsoidal of varying volume, shape, and orientation).

Probability of cluster membership **K** (p*_ik_*) is then normalized relative to all clusters 1 to *K* in the Expectation step*.*

Proportion (π*_k_*), mean (μ*_k_*), and covariance (Σ*_k_*) of cluster k are then updated based on the above updated probability in the Maximization step.

We identified the optimal number of clusters through bootstrap resampling and evaluation of the adjusted Rand Index, separately for both the VEV and VVV specifications. Both models suggested an optimal number of clusters **K** of 5. We chose the VVV specification given its more flexible nature.
